# DISTINCT SALIVARY MICROBIOTA PROFILES, *BDNF* DNA METHYLATION AND *MIR-16-5P*, *MIR-29A-3P*, *MIR-191-5P* ALTERATIONS IN OBSESSIVE-COMPULSIVE DISORDER AND MAJOR DEPRESSIVE DISORDER

**DOI:** 10.1101/2025.01.24.25321067

**Authors:** Antonio Girella, Matteo Vismara, Kenneth J. O’Riordan, Eoin Gunnigle, Francesca Mercante, Nicolaja Girone, Mariangela Pucci, Valentina Gatta, Fani Konstantinidou, Liborio Stuppia, John F. Cryan, Bernardo Dell’Osso, Claudio D’Addario

## Abstract

Obsessive-Compulsive Disorder (OCD) and Major Depressive Disorder (MDD) frequently co-occur, with depressive symptoms affecting OCD progression and vice versa. Identifying biomarkers is crucial for improving diagnosis and treatment. While the gut microbiota’s role in psychiatric disorders is well-studied, this research focuses on alterations in the oral microbiota and their relationship with *BDNF* (Brain-Derived Neurotrophic Factor) DNA methylation in OCD and MDD patients compared to healthy controls. Our findings reveal significant changes in microbiota composition with OCD patients showing increased Actinobacteriota and Firmicutes abundances (*p<0.05*; CTRL=n.24, OCD=n.21), while MDD patients exhibiting increased Actinobacteriota and Firmicutes, with reduced Bacteroidota and Proteobacteria abundances (*p<0.05*; CTRL=n.24, MDD=n.16). These alterations, including potential post-streptococcal autoimmunity, highlight the microbiota’s role in OCD and MDD pathophysiology. Selective changes in *BDNF* DNA methylation were observed in both disorders at CpG sites in exon I and IV, significantly reduced in OCD and MDD (*p<0.05*; CTRL=n.24, OCD=n.23, MDD=n.16) and, following miRNome analysis showed altered expression of *BDNF*-targeting microRNAs, with *miR-16-5p* and *miR-29a-3p* upregulated in OCD (*p<0.05*; CTRL=n.24, OCD=n.17), and *miR-29a-3p* upregulated and *miR-191-5p* downregulated in MDD (*p<0.05*; CTRL=n.24, MDD=n.16). These findings suggest disorder-specific microbiota and epigenetic profiles, positioning saliva as a non-invasive tool for biomarker identification. This research advances understanding of microbial-epigenetic interactions in OCD and MDD, potentially guiding early diagnosis and targeted therapies.

## Introduction

Obsessive-Compulsive Disorder (OCD) and Major Depressive Disorder (MDD) are two common and debilitating psychiatric disorders [1–3]. Although classified as separate conditions, MDD is frequently reported as a comorbidity in individuals with OCD. The severity of OCD symptoms and their profound impact on daily functioning can contribute to the development of MDD and additionally, depressive symptoms may alter the course of OCD [4]. On the other hand, obsessive and compulsive symptoms are often observed in severe cases of MDD, which is also indicated by scales such as the Hamilton Depression Rating Scale where these symptoms are included in supplementary items to assess the severity of depression [5]. It has been also observed that, among patients suffering from OCD, 65-85% manifest other psychiatric conditions, with the most common comorbid diagnoses are anxiety disorders [6, 7], mood disorders [6, 7], and in particular with major depressive disorder [6, 8, 9] OCD shows high genetic correlations with [10, 11]. Moreover, OCD and MDD share not only common psychopathologic characteristics but also treatment response to serotoninergic medications. Prospective studies on OCD and MDD comorbidity indicate that frequently OCD symptoms anticipate depressive symptoms, as mood changes may occur as a response to the chronic distress and functional impairment associated with the burden of OCD [12].

The complexity of their pathological mechanisms makes pharmacological, psychotherapeutic, or somatic treatments not satisfactory for people suffering from these two mental disorders, with a worsened quality of life and higher risk of suicide [2, 13].

It is thus of relevance to better understand the mechanisms underlying their development and identify potential biomarkers to gain a more accurate and speedy diagnosis and improve treatment response. As with many mental health issues, MDD and OCD are multifactorial and polygenic disorders, where the interplay between genes and environmental factors, internal and external, is relevant [14–17].

Over the last years, many studies have focused on the role of the gut microbiota in the pathophysiology of psychiatric disorders, including MDD and OCD [18–20]. It is becoming increasingly accepted that the gut microbiota is susceptible to several internal and external factors, including genetic, diet, lifestyle, aging, antibiotics use, and mode of delivery [21]. Although the gut-microbiota has been widely studied for its role in health and disease, the microbiota present in the oral cavity is becoming increasingly popular, since it reflects the diversity and composition of that in the gastrointestinal tract [22]. Recent studies have focused on the impact of oral microbiota in psychiatric disorders; however, its role remains under characterized [23, 24]. It has been shown that oral microbiome health is not only an indicator of oral health problems but is also a key player in systemic disease, including obesity, diabetes, and neurological and psychiatric disorders including OCD and MDD [25, 26].

The microbiota can affect host physiology acting on epigenetic regulation through different mechanisms. Bacterial metabolite production drive epigenetic modulation, regulate the expression and activity of key epigenetic enzymes or activate host-cell processes acting on epigenetic pathways [27].

We and others have already reported the epigenetic regulation of several genes in both disorders [28, 29] and, among others, one of the most studied is *Brain-Derived Neurotrophic Factor* (*BDNF*). Human *BDNF* gene is quite complex and formed by 11 exons (I-IX, Vh and VIIIh) and 9 promoters [30]. *BDNF* promoter exon I (*BDNF*-I) and IV (*BDNF*-IV) DNA methylation was already investigated in MDD [31, 32] and OCD [33, 34]. In particular, higher levels of DNA methylation at *BDNF* gene in peripheral blood mononucleated cells (PBMCs) were reported in patients suffering from MDD when compared to healthy controls [31, 32], on the other hand an hypomethylation occurred at specific CpG sites on *BDNF* exon I [35] and exon IV [34, 35]in blood samples of MDD individuals versus controls. Alterations of *BDNF* methylation pattern have been also observed in both blood and saliva samples of OCD patients compared to healthy controls, namely a reduction in the methylation levels of *BDNF* gene at exon I promoter region accompanied by an increase in the expression level of the gene [33, 34]. However, an increase in peripheral blood *BDNF* expression level in non-medicated individual suffering from OCD has been reported too [37].

Circulating microRNAs (miRNAs) have also been suggested as potential biomarkers of OCD [38, 39] and MDD [40, 41] and among these miRNAs that directly target *BDNF* have been associated with psychiatric conditions including depression [42–51]. Circulating miRNAs are a type of regulatory non-coding RNA often incorporated into exosomes as signaling molecules making them potential candidates for mental health biomarker discovery, for both prognosis and diagnosis of disease, as well as treatment response [52, 53]. Moreover, studies have reported how metabolites produced by the gut microbiota might influence the expression of host miRNAs [54, 55], as well as how host miRNAs might influence gut microbiota composition [56]. We investigated the possible modulation of exosomal miRNAs in OCD and MDD. miRNome analysis showed 126 miRNAs differentially modulated in OCD and MDD patients compared to CTRLs. Among these 126 miRNAs, we chose to analyze 4 of these targeting *BDNF* gene and known to be involved in the pathogenesis and development of different psychiatric disorders. *miR-16-5p* for its role in regulating BDNF has already reported to be involved in depressive-like behaviors induced by early-life stress [56], in schizophrenia [42], as well as *in vitro* in SH-SY5Y growth [57]. *miR-29a-3p* was found significantly upregulated in depressed rats and in patients suffering from autism spectrum disorder [59, 60]; *miR-191-5p,* involved in long-lasting depression [61, 62]; *miR-202-3p* resulted to be upregulated in the hippocampus of rats showing depressive-like behaviors [63].

Here we investigated if alterations in the salivary microbiome composition might occur in OCD and MDD and if epigenetic regulation might account for it. Using saliva samples collected from MDD and OCD subjects we thus analyzed (1) microbiome diversity and composition; (2) *BDNF* DNA methylation levels; (3) exosomal miRNAs levels; (4) correlations between the molecular markers and the salivary microbiome.

Finally, since microbiota can modulate brain function and development [64], and it has been shown that repetitive transcranial magnetic stimulation (TMS) is an effective and noninvasive treatment for both MDD [65] and OCD [66], we also aimed to investigate the recently suggested interplay between TMS and microbiota composition [67] in a subset of MDD and OCD subjects.

## Materials and Methods

### Participants

Patients with OCD or MDD were recruited at the outpatient psychiatric clinic at Sacco University Hospital in Milan, Italy. Diagnoses were assessed by psychiatrists trained in the diagnosis and treatment of these conditions, through the administration of a semi-structured interview based on DSM-5 criteria (SCID 5 research version, RV) [68]. In case of psychiatric comorbidity, OCD or MDD had to be the primary disorder. A total of 23 patients suffering from OCD (13 women and 10 men; age: 40.82 ± 12.00) and 16 patients suffering from MDD (11 women and 5 men; age: 53.50 ± 13.85) were included in the study. The following socio-demographic and clinical characteristics were collected: age, gender, marital status, employment, educational status, psychiatric family history, age at OCD/MDD onset, psychiatric and medical comorbidities, duration of illness, duration of untreated illness (DUI), age at first treatment, and current pharmacological and psychological treatments. Illness severity was measured through the Yale-Brown Obsessive– Compulsive Scale in patients with OCD [69] and with the Hamilton Depression Rating Scale (HAM-D) and the Montgomery-Asberg Depression Rating Scale (MADRS) in patients with MDD [70, 71]. The YBOCS and the HAM-D were administered at baseline and at the end of the TMS stimulation, to stratify patients according to response to treatment. In detail, OCD patients were considered responders if they showed a YBOCS score reduction ≥ 35%, while MDD patients were considered responders if they showed a HAM-D score reduction ≥ 25%; alternatively, in both subgroups, patients were considered non-responders. Twenty-four healthy control (CTRL) volunteers were recruited (13 women and 11 men; age: 41.50 ± 11.97) from University of Teramo, Italy. Controls were matched for age and sex, without any psychiatric disorder, as determined by the non-patient edition of the SCID and no positive family history for major psychiatric disorders in the first-degree relatives [72]. All participants had given their written informed consent to participate in the study, which included the use of personal and clinical data as well as saliva drawing for the analysis. The study protocol had been previously approved by the local Ethics Committee. Demographic and clinical characteristics for the OCD and MDD subjects are shown in Table 1 and in Supplementary Table 1.

**Table 1.** Socio-demographic and clinical features of CTRL group, MDD and OCD patients. Categorical variables have been analyzed by Chi-squared test (χ2), while continuous variables have been analyzed basing on normality distribution test with parametric (Welch’s test) or nonparametric (Mann-Whitney test, for two groups or Kruskal-Wallis test, for three groups) t-test. Legend: BD, bipolar disorders; FED: feeding and eating disorders; GAD: general anxiety disorders; PD: panic disorder.

| ***SOCIO-DEMOGRAPHIC FEATURES*** | **CTRL (n=24)** | **OCD (n=23)** | **MDD (n=16)** | **P value** |
| --- | --- | --- | --- | --- |
| Gender (%) | 45.8 (male)<br>54.2 (female) | 43.5 (male)<br>56.5 (female) | 31.2 (male)<br>68.8 (female) | 0.633 ($\chi^2$) |
| Age at recruitment (mean ± SD) | 41.5 ± 12.0 | 40.1 ± 11.6 | 53.5 ± 14.1 | 0.008 (Kruskal-Wallis test) |
| Education: graduated (%) | 95.8 | 21.8 | 31.5 | < 0.0001 ($\chi^2$) |
| Employment: employed (%) | 100 | 78.2 | 81.2 | 0.057 ($\chi^2$) |
| Married/engaged (%) | 83.3 | 30.4 | 68.8 | < 0.001 ($\chi^2$) |
| ***CLINICAL FEATURES*** |  |  |  |  |
| Age at onset (years, mean ± SD) | - | 21.7 ± 9.8 | 32.4 ± 10.4 | < 0.001 (nonparametric t-test) |
| Family history of psychiatric disorder (%) | - | 65.2 | 81.3 | 0.274 ($\chi^2$) |
| Psychiatric comorbidity (%) | - | 78.3 | 56.3 | 0.143 ($\chi^2$) |
| Affective disorders (MDD, BD) | - | 16.7 | - | - |
| Anxiety disorders | - | 77.8 | 25 | 0.001 ($\chi^2$) |
| (PD, GAD) |  |  |  |  |
| OCD | - | - | 12.5 | - |
| Personality disorders | - | - | 6.3 | - |
| Substance use disorders | - | - | 18.8 | - |
| FED | | 5.6 | 6.3 | 0.702 ($\chi^2$) |
| Medical comorbidity (%) | - | 30.4 | 43.8 | 0.571 ($\chi^2$) |
| Duration of illness (years, mean ± SD) | - | 18.4 ± 12.4 | 20.4 ± 10.2 | 0.584 (parametric t-test) |
| **Psychometric variables** |  |  |  |  |
| Y-BOCS score at assesment (mean ± SD) | - | 24.3 ± 7 | - | - |
| HAMD score at assesment (mean ± SD) | - | - | 17.3 ± 6.1 | - |
| MADRS score at assesment (mean ± SD) | - | - | 25.01 ± 7.6 | - |
| **Current treatment (%)** |  |  |  |  |
| Antidepressants | - | 91 | 100 | 0.226 ($\chi^2$) |
| Mood stabilizers | - | 13 | 37.5 | 0.075 ($\chi^2$) |
| Antipsychotics | - | 43.5 | 68.8 | 0.119 ($\chi^2$) |
| Benzodiazepines | - | 30.4 | 56.3 | 0.107 ($\chi^2$) |
| Drug-free | - | 4.3 | 0 | 0.398 ($\chi^2$) |
| Psychoterapy | - | 29.1 | 43.8 | 0.571 ($\chi^2$) |
| **TMS treatment responsiveness (%)** | - | 30.0 | 63.5 | 0.107 ($\chi^2$) |

### TMS stimulation

All MDD patients and 10 of the 23 OCD patients underwent repetitive TMS treatment. For more details, please refer to Supplementary information.

### Sampling

Saliva samples were collected from OCD and MDD patients as well as from the CTRL group in the morning. For those who underwent TMS, saliva was collected twice, before and after the treatment. Donors were required to abstain from eating, drinking, smoking, or brushing their teeth for at least one hour prior to collection. Approximately 2 mL of saliva from each individual were collected in 15 mL tube and stored at −80°C until processing.

### DNA extraction

DNA was extracted from saliva samples through salting out method following Garbieri and collaborators protocol [73], followed by an additional cleaning step using the DNA Clean & Concentrator-5 kit (Zymo Research, Orange, CA, USA) according to the manufacturer’s protocol.

### 16S rRNA Microbiome sequencing

DNA was diluted to 5 ng/ul and the V3-V4 hypervariable region of the 16S rRNA gene was amplified from the DNA extracts and prepared for sequencing using the Illumina 16S Metagenomic Sequencing Library Protocol (http://www.illumina.com/content/dam/illumina-support/documents/documentation/chemistry_documentation/16s/16s-metagenomic-library-prep-guide-15044223-b.pdf). Final libraries were quantified and pooled equimolarly and the concentration of the final pool was determined using the Qubit high sensitivity assay (Life Technologies). The final pool was also run on an Agilent Bioanalyser high Sensitivity Chip (Agilent) to assess the quality of the final pool. The pool was then diluted, combined with PhiX (40%), and sequenced on a NextSeq 2000 using a P1 600 cycle reagent kit according to manufacturer’s instructions (Illumina).

### DNA methylation analysis by Pyrosequencing

Bisulfite converted genomic DNA (500 ng) was amplified with a biotin-labelled primer using the PyroMark PCR kit (Qiagen, Hilden, Germany) in accordance with manufacturer’s guidelines. Pyrosequencing was performed by a PyroMark Q48 Autoprep using PyroMark Q48 Advanced Reagents (Qiagen) as already described in our previous works [74, 75]. Primers used, sequences analyzed, and genome localization are reported in Supplementary Table 2.

### Exosome isolation and miRNAs extraction

Exosomes were extracted from 1 mL of saliva using Total Exosome Isolation Reagent (Invitrogen by Thermo Fisher Scientific, Vilnius, Lithuania). miRNAs were isolated from exosomes through Total Exosome RNA and protein isolation kit (Invitrogen by Thermo Fisher Scientific) according to the manufacturer’s protocol.

### miRNA analysis

Exosomal RNA belonging to all participants were pooled in 3 groups (CTRL group, OCD group and MDD group). Exosomal RNA was quantified through Invitrogen Qubit 4 Fluorometer (CTRL pool = 42.1 ng/µl, 260/280 = 1.69, 260/230 = 1.82; OCD pool = 19.7 ng/ µl, 260/280 = 1.76, 260/230 = 1.77; MDD pool = 11.9 ng/µl, 260/280 = 1.70, 260/230 = 1.69) and miRNome analysis was performed. TaqMan™ Array Human MicroRNA A Cards v2.0 (Applied Biosystems, Foster City, CA, USA) containing 384 miRNAs was used. Then, complementary cDNA was synthetised by miRCURY LNA RT kit (Qiagen) following manufacturer’s recommendations and four miRNAs, selected from miRNome analysis, were quantified through quantitative real-time PCR (qRT-PCR) using SensiFAST SYBR Lo-ROX Kit (Bioline Reagents, London, UK) on QIAquant 96 5plex (Qiagen). Both miRNome and single miRNAs results were first normalized to two reference genes *U6* and *U48*, then differences between groups were calculated by the Delta-Delta Ct (ΔΔCt) method and converted to fold change expressed as 2^(−ΔΔCt)^ [76]. Sequences used for amplification are listed in Supplementary Table 3.

### Statistical analysis

Socio-demographic and clinical features of CTRL group, MDD and OCD patients’ groups have been analyzed. Namely, categorical variables have been analyzed by Chi-squared test, while continuous variables have been analyzed basing on normality distribution test (normal distribution: Welch’s t-test; not normal distribution: Mann-Whitney test (two groups) and Kruskal-Wallis test with Dunn’s correction for multiple comparison (three groups)). Statistical analysis was initially performed by treating pre- and post-TMS samples as independent data sets. Subsequently, additional analyses were conducted to evaluate the effects of TMS by comparing pre- and post-TMS measurements within the same individuals. Statistical differences in DNA methylation and in miRNAs expression were determined using GraphPad Prism 9 (Graph-Pad Software, San Diego, CA). Normality tests were conducted. For DNA methylation data distribution resulted to be normal for both OCD and MDD groups when comparing to CTRL group, even stratifying for pre- and post-TMS treatment (Shapiro-Wilk *p* > 0.05 and Kolmogorov-Smirnov *p* > 0.1). While, for miRNAs analysis the distribution resulted to be not normal for all miRNAs analyzed, even stratifying for pre- and post-TMS treatment (Shapiro-Wilk *p* < 0.05 and Kolmogorov-Smirnov *p* < 0.1). DNA methylation at each CpG site was analyzed using multiple unpaired t-test and False Discovery Rate (FDR) 10% Benjamini-Hochberg (BH) correction was used for the multiple comparisons between the experimental groups with a q-value of 0.1 used as a cut-off. While Two-way ANOVA with FDR 10% BH correction (0.1 used as cut-off) was used for multiple comparisons between three groups. Single miRNAs were analyzed using Mann-Whitney test. When considering TMS effects comparing pre- and post-TMS, single miRNAs were analyzed with Kruskal-Wallis test with Dunn’s correction for the multiple comparison between three groups. The statistical power of the epigenetic analyses was evaluated using G*Power 3.1. With the study’s sample size and assuming a medium effect size (f = 0.25) and an alpha of 0.05, the power exceeded 0.80, ensuring adequate sensitivity to detect significant differences.

For microbiome analysis, paired-end reads were pre-filtered based on a quality score threshold of > 28, trimmed and filtered for quality and chimaeras using the DADA2 library [77] in R (version 4.3.2). Only samples with > 10,000 reads after quality control steps were used in the analysis. Taxonomy was assigned using the DADA2 package that included the SILVA SSURef database release v138.1 [78]. Parameters as recommended in the DADA2 manual were adhered to unless mentioned otherwise. ASVs were aggregated at genus level; those that were unknown at the genus level were not considered in downstream analysis. Further data-handling was undertaken in R (version 4.3.2) using Rstudio GUI (version 1.1.453). Principal component analysis was performed on centred log-ratio transformed (clr) values using the ALDEx2 library [79]. The number of permutations was always set to 1000. Beta diversity was computed in terms of Aitchison distance, or Euclidean distance between CLR-transformed data. Alpha diversity was computed using the iNEXT library [80]. Stacked barplots were generated by normalising counts to 1, generating proportions. Genera that were never detected at a 2% relative abundance or higher were aggregated and defined as rare taxa for the purposes of the stacked barplots. Differential abundance of microbes were calculated using implementations of the ALDEx2 library. A *p* value of < 0.05 was deemed significant in all cases. To correct for multiple testing in tests involving microbiota or functional modules, the Benjamini-Hochberg (BH) post-hoc was performed with a q-value of 0.1 used as a cut-off. Custom R scripts and functions are available at https://github.com/thomazbastiaanssen/Tjazi.

All the data were compared by Spearman’s rank correlation coefficient. The *p* values < 0.05 were statistically significant.

## Results

The study included 24 controls (CTRL), 23 OCD patients, and 16 MDD patients. Gender distribution was comparable across groups (*p* = 0.633), with females representing 54.2%, 56.5%, and 68.8% in CTRL, OCD, and MDD groups, respectively. The mean age at recruitment was significantly higher in the MDD group (53.5 ± 14.1 years) compared to CTRL (41.5 ± 12.0 years) and OCD (40.1 ± 11.6 years) (*p* = 0.008). Educational attainment differed significantly (*p* < 0.0001), with 95.8% of CTRL participants being graduates, compared to 21.8% in OCD and 31.5% in MDD groups. Employment rates were high in all groups but lower in OCD (78.2%) compared to CTRL (100%) and MDD (81.2%) (*p* = 0.057). Marital status showed significant variation, with married/engaged participants comprising 83.3% in CTRL, 30.4% in OCD, and 68.8% in MDD groups (*p* < 0.001). Clinical features highlighted distinct patterns between groups, particularly for age at onset and comorbidities, detailed in Table 1. The study also included 10 OCD patients and 16 MDD patients who underwent TMS treatment. Baseline psychometric assessments showed a mean Y-BOCS score of 25.0 ± 8.42 for OCD and mean HAMD and MADRS scores of 17.7 ± 4.71 and 24.6 ± 5.54, respectively, for MDD. Post-TMS assessments indicated reductions in scores across both groups, with a non-significant decrease in Y-BOCS for OCD (*p* = 0.072) and a significant reduction in HAMD for MDD (*p* = 0.022). Changes in MADRS were not significant (*p* = 0.137). Results highlight the distinct psychometric profiles and response patterns of OCD and MDD patients. For further details, refer to Supplementary Table 1.

### Microbiota analysis

We reported microbial alpha diversity indices in Figure 1A-C. We observed significant differences in alpha diversity indices between OCD patients and CTRLs, for species evenness: Simpson Index (*p* = 0.035, Tukey-HSD correction) and Shannon Entropy (*p* = 0.027, Tukey-HSD correction) (Figure 1B, C), whereas no statistically significant differences are shown regarding species richness (Chao1, *p* = 0.161, Tukey-HSD correction) (Figure 1A). There were no significant differences in species evenness considering microbiota composition in saliva in MDD patients when compared to CTRLs (Figure 1B, C, Tukey-HSD correction). Beta diversity was evaluated through PCA analysis showing a statistically significant community dissimilarity between the groups under study. Specifically, significant difference was shown considering MDD patients’ microbiota composition compared to CTRL individuals (*p* = 0.001), but no significant difference resulted from the comparison between OCD group and CTRLs (*p* = 0.153) (Figure 1D). However, stratifying both OCD and MDD patients based on TMS treatment, no significant differences in both richness (Chao1) and evenness (Simpson Index and Shannon Entropy) considering pre and post TMS treatment (Supplementary Figure 2A, B, C) was observed, whereas there is a significant community dissimilarity in beta diversity index (*p* = 0.001) mainly in the MDD group (Supplementary Figure 2D).

**Figure 1.**
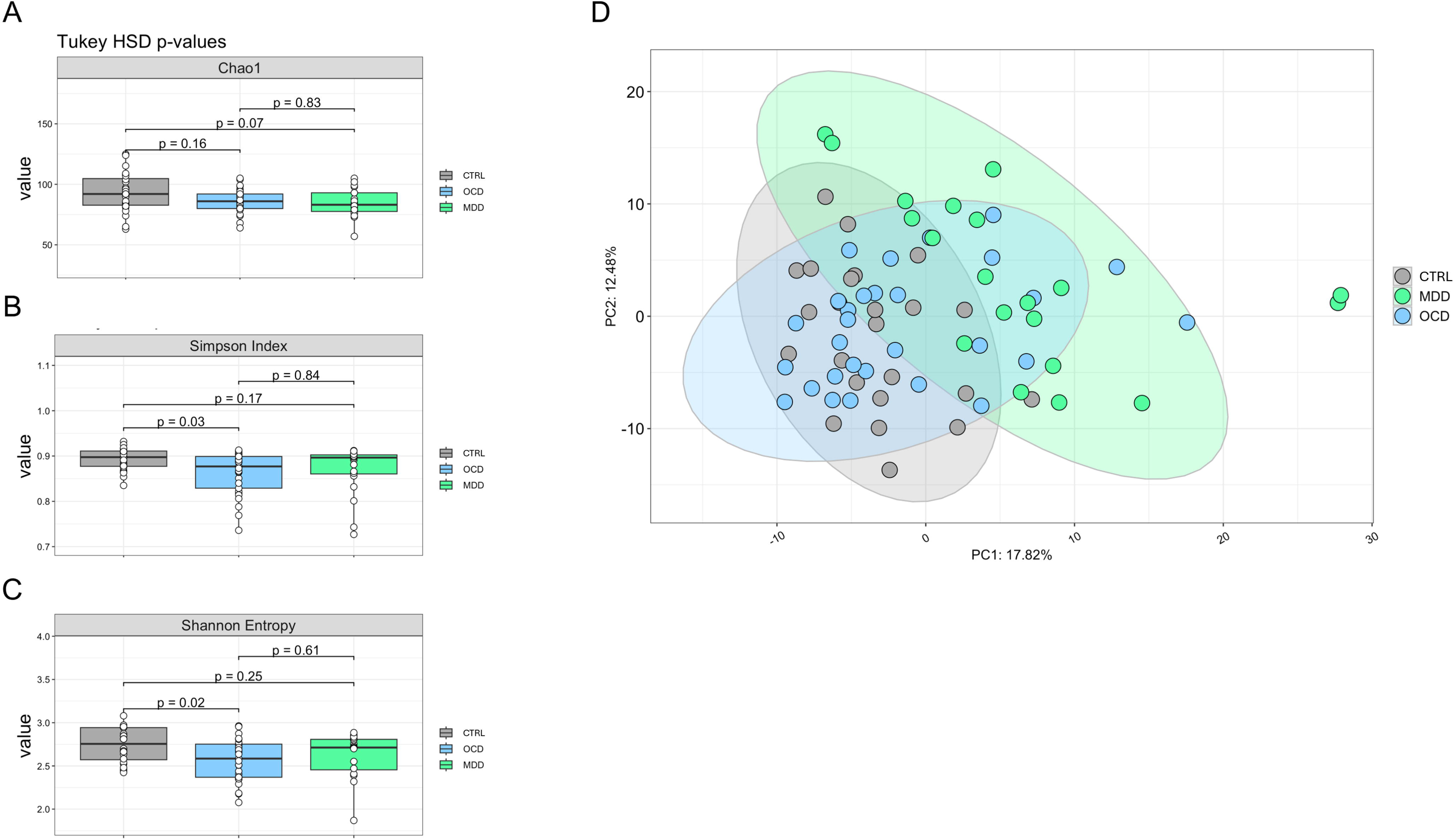
Salivary microbiota composition in CTRL healthy individuals (n=24), OCD (n=29) and MDD patients (n=20). Alpha diversity indices: (**A**) Chao1, (**B**) Simpson index and (**C**) Shannon entropy. (**D**) PCA plot of beta diversity of salivary microbiota of CTRLs, OCD and MDD patients. Data considered to be significant with a *p* value < 0.05 (Tukey-Honestly Significant Difference (HSD)).

Analyses for each taxon abundance considering OCD, MDD patients and CTRLs (Supplementary Figure 1) revealed, among 246 taxa identified, 7 phyla (Actinobacteriota, Bacteroidota, Campylobacterota, Firmicutes, Fusobacteriota, Proteobacteria, Synergistota) and 11 bacterial genera (*Rothia*, *Alloprevotella*, *Campylobacter*, *Streptococcus*, *Gemella*, *Catonella*, *W5053*, *Sneathia*, *Simonsiella*, *Enhydrobacter*, *Pyramidobacter*; 4.4%, out 246 in total) significantly altered in OCD patients compared to the CTRL group (*p* < 0.05, Tukey-GLM 10% FDR correction), in specific *Rothia*, *Streptococcus*, *Gemella*, *W5053*, *Sneathia*, *Enhydrobacter*, *Pyramidobacter* resulted to be more abundant, and *Alloprevotella*, *Campylobacter*, *Catonella*, *Simonsiella* were less abundant in saliva from OCD patients with respect to CTRLs (Figure 2A). In MDD we found to be differentially abundant in patients when compared to CTRLs, 7 phyla (Actinobacteriota, Bacteroidota, Campylobacterota, Desulfobacterota, Firmicutes, Fusobacteriota, Proteobacteria) and 38 genera (15%, out 246 in total) among which 19 genera (*Gardnerella*, *Scardovia*, *Rothia*, *Olsenella*, *Cryptobacterium*, *Slackia*, *Prevotellaceae UCG-004*, *Desulfobulbus*, *Asteroplasma*, *Lacticaseibacillus*, *Lactobacillus*, *Limosilactobacillus*, *Pseudoramibacter*, *Howardella*, *Shuttleworthia*, *Parvimonas*, *Peptoniphilus*, *Selenomonas*, *Dialister*) more abundant, and 19 less abundant in patients (*Pseudopropionibacterium*, *F0058*, *Alloprevotella*, *Bergeyella*, *Campylobacter*, *Catonella*, *Jonhsonella*, *Ruminococcus*, *Peptococcus*, *Eubacterium yurii*, *Oceanvirga*, *Lautropia*, *Alysiella*, *Kingella*, *Neisseria*, *Simonsiella*, *Actinobacillus*, *Aggregatibacter*, *Haemophilus*) (*p* < 0.05, Tukey-GLM 10% FDR correction) (Figure 2B). We also investigated microbiota composition before and after TMS treatment in both OCD and MDD patients (Supplementary Figure 3). We reported that 2 genera out of 246 identified in total (0.8%) (Supplementary Figure 4A) were differently modulated in saliva samples from post-TMS OCD patients. In particular, *Gemella* and *Streptococcus* (Firmicutes) abundances were significantly increased compared to CTRLs (*p* < 0.05, Tukey-GLM 10% FDR correction). In MDD patients we showed no significant modulations in taxa abundances driven by TMS treatment (Supplementary Figure 4B, C), except for *Catonella* (Firmicutes) which was significantly decreased in the pre-TMS MDD group (*p* < 0.05, Tukey-GLM 10% FDR correction), while in post-TMS MDD group returned to the levels of the CTRL group (Supplementary Figure 4C).

**Figure 2.**
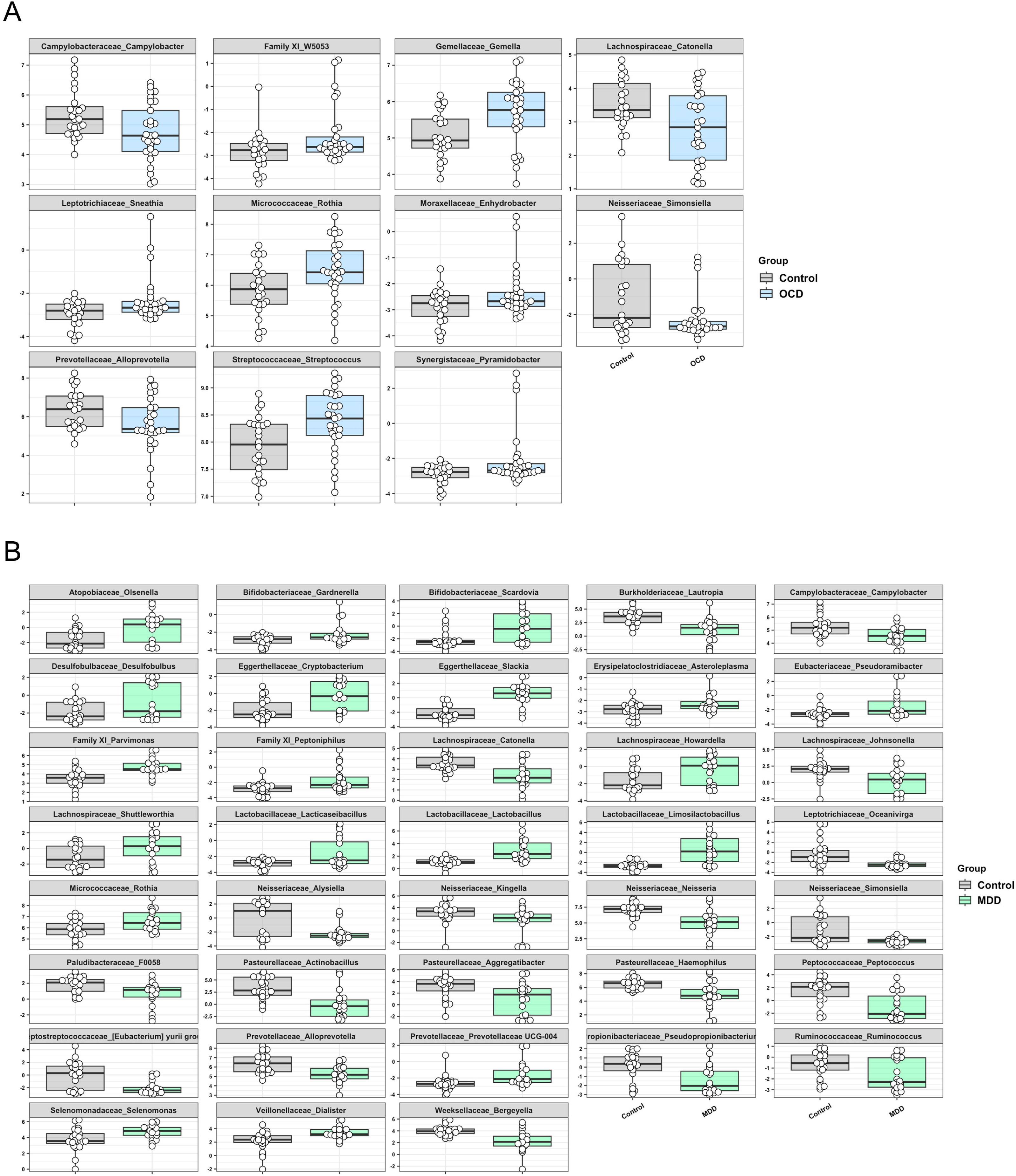
Salivary microbiota composition in CTRL healthy individuals (n=24), OCD (n=29) and MDD patients (n=20). (**A**) Differentially abundant genera from saliva of CTRL vs OCD individuals. (**B**) Differentially abundant genera from saliva of CTRL vs MDD individuals. Data considered to be significant with an adjusted *p* value < 0.05 (Tuckey-Generalised Linear Model (GLM) 10% FDR correction).

### *BDNF* gene methylation

Methylation pattern of two different CpG islands marking two different promoter regions among *BDNF* gene, namely *BDNF*-I and *BDNF*-IV has been analyzed (Figure 3A). Regarding *BDNF*-I, here we report a significant reduction of methylation levels when comparing OCD patients with CTRLs on CpG site 3 (OCD: 2.78 ± 0.61; CTRL: 3.97 ± 2.31; *p* = 0.024, corrected q = 0.061) and considering the average (AVE) of all CpG sites under study (OCD: 3.01 ± 0.57; CTRL: 4.04 ± 1.75; *p* = 0.012, corrected q = 0.061) (Figure 3B). A significant hypomethylation also occurred in saliva samples from OCD patients compared with CTRLs at CpG site 2 (OCD: 4.59 ± 1.12; CTRL: 6.40 ± 1.17; *p* < 0.0001, corrected q < 0.0001), site 3 (OCD: 3.87 ± 0.89; CTRL: 5.05 ± 0.79; *p* < 0.0001, corrected q < 0.0001), site 4 (OCD: 3.48 ± 0.52; CTRL: 4.12 ± 0.97; *p* < 0.01, corrected q = 0.011), site 5 (OCD: 4.64 ± 1.06; CTRL: 5.95 ± 1.33; *p* < 0.001, corrected q < 0.001) as well as at the average of all sites (OCD: 3.47 ± 0.42; CTRL: 4.35 ± 0.71; *p* < 0.0001, corrected q < 0.0001) for BDNF-IV (Figure 3C). Stratifying data by TMS there were no differences considering *BDNF*-I (Supplementary Figure 5A), while TMS induced a slight effect on the epigenetic mark at *BDNF*-IV, decreasing the significance between CTRL group and OCD patients, except for site 2 (CTRL: 6.39 ± 1.17; pre-TMS OCD: 4.27 ± 0.62; *p* < 0.0001, corrected q < 0.0001; post-TMS OCD: 4.49 ± 0.64; *p* < 0.0001, corrected q < 0.0001); site 3 (CTRL: 5.05 ± 0.79; pre-TMS OCD: 3.23 ± 0.62; *p* < 0.0001, corrected q < 0.0001; post-TMS OCD: 3.86 ± 0.65; *p* < 0.001, corrected q < 0.001); site 4 (CTRL: 4.12 ± 0.97; pre-TMS OCD: 3.17 ± 0.53; *p* = 0.006, corrected q = 0.002; post-TMS OCD: 3.58 ± 0.56; *p* > 0.05, no significant); site 5 (CTRL: 5.95 ± 1.33; pre-TMS OCD: 4.47 ± 0.84; *p* < 0.0001, corrected q < 0.0001; post-TMS OCD: 5.06 ± 0.82; *p* = 0.006, corrected q = 0.004); AVE (CTRL: 4.35 ± 0.71; pre-TMS OCD: 3.17 ± 0.29; *p* < 0.001, corrected q < 0.001; post-TMS OCD: 3.34 ± 0.34; *p* = 0.001, corrected q = 0.001) (Supplementary Figure 5B). Moreover, we saw no significant differences considering patients’ response to TMS treatment (Supplementary Figure 6A).

**Figure 3.**
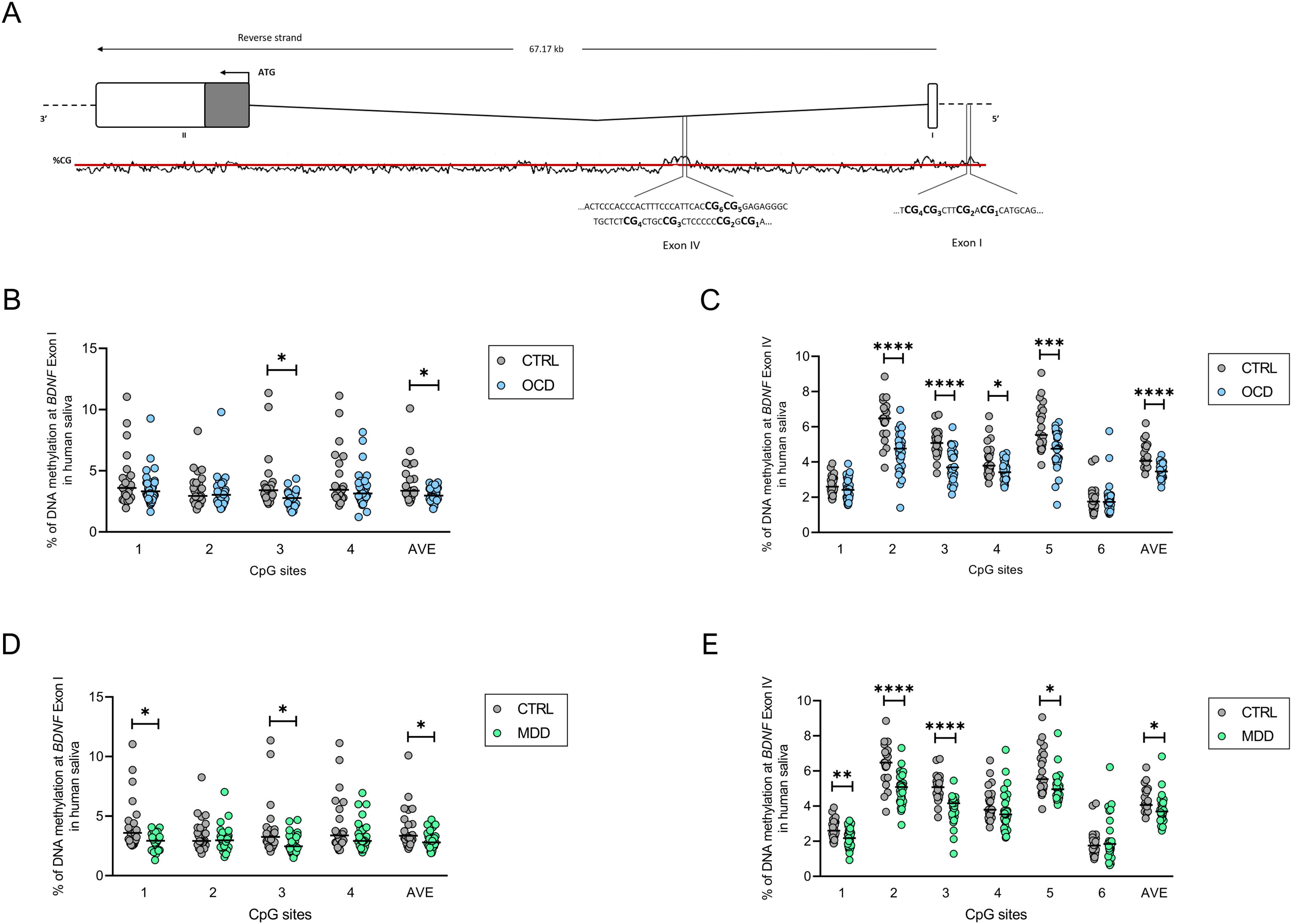
*BDNF* gene regulation in saliva samples. (**A**) Schematic representation of human *BDNF* gene (Transcript BDNF-201; ENST00000314915.6, genome assembly GRCh38:CM000673.2). ATG is the translation start site: filled boxes represent the exons’ translated sequence. In the lower part is shown the CpG islands with their sequences and the position of the CpG sites under study. % of DNA methylation at *BDNF* exon I promoter (4 CpG sites) considering CTRLs vs OCD (**B**) and CTRLs vs MDD (**D**). % of DNA methylation at *BDNF* exon IV promoter (6 CpG sites) considering CTRLs vs OCD (**C**) and CTRLs vs MDD (**E**). (Unpaired t-test and FDR 10% BH correction, q < 0.1 considered to be significant; CTRL n=24; OCD n=29; MDD n=20). Data are represented as scatter dots with medians.

The same pattern of changes has been observed in saliva obtained from individuals suffering from MDD when compared to CTRL individuals. In particular, DNA from MDD subjects was hypomethylated compared to CTRLs at *BDNF*-I promoter region considering CpG site 1 (MDD: 2.83 ± 0.75; CTRL: 4.32 ± 2.22; *p* < 0.01, corrected q = 0.026), CpG site 3 (MDD: 2.70 ± 0.87; CTRL: 3.89 ± 2.30; *p* = 0.024, corrected q = 0.041) and also the AVE of all sites (MDD: 2.95 ± 0.74; CTRL: 3.96 ± 1.75; *p* = 0.012, corrected q = 0.031) (Figure 3D). A significant reduction in the percentage of DNA methylation in MDD patients has also been reported at CpG sites 1 (MDD: 2.17 ± 0.57; CTRL: 2.70 ± 0.57; *p* < 0.01, corrected q < 0.01), 2 (MDD: 4.96 ± 0.90; CTRL: 6.40 ± 1.17; *p* < 0.0001, corrected q < 0.0001), 3 (MDD: 3.88 ± 0.81; CTRL: 5.05 ± 0.79; *p* < 0.0001, corrected q < 0.0001), 5 (MDD: 5.08 ± 0.86; CTRL: 5.95 ± 1.33; *p* = 0.012, corrected q = 0.021), and considering all sites’ average (MDD: 3.83 ± 0.80; CTRL: 4.36 ± 0.71; *p* = 0.016, corrected q = 0.023) at exon IV promoter region (Figure 3E).

Although, we do not report any significant differences in DNA methylation considering TMS treatment at *BDNF*-I in saliva from MDD individuals (Supplementary Figure 5C), a statistically significant decrease in methylation pattern of *BDNF*-IV was noticed in post-TMS MDD group: site 2 (CTRL: 6.39 ± 1.17; pre-TMS MDD: 4.70 ± 0.73; *p* < 0.0001, corrected q < 0.0001; post-TMS MDD: 5.20 ± 0.99; *p* < 0.001, corrected q < 0.001); site 3 (CTRL: 5.05 ± 0.79; pre-TMS MDD: 3.66 ± 0.95; *p* < 0.001, corrected q < 0.001; post-TMS MDD: 4.09 ± 0.60; *p* = 0.008, corrected q = 0.005) and AVE (CTRL: 4.40 ± 0.69; pre-TMS MDD: 3.49 ± 0.55; *p* = 0.041, corrected q = 0.014; post-TMS MDD: 4.12 ± 1.05; *p* > 0.05, no significant) (Supplementary Figure 5D). We also saw a significant increase in the percentage of methylation at *BDNF*-IV site 2 considering the response after TMS treatment (no responsive: 4.41 ± 0.26; responsive: 6.02 ± 0.66; *p* < 0.001, corrected q = 0.003) (Supplementary Figure 6B). Moreover, stratifying *BDNF* methylation data for the pharmacotherapy within OCD and MDD patients, in terms of monotherapy (antidepressant or antipsychotic/mood stabilizer/benzodiazepines) and multitherapy (two or more of those mentioned) we show significant alteration in OCD for *BDNF*-I site 1 (monotherapy: 2.82 ± 0.66; multitherapy: 3.62 ± 0.98; *p* = 0.025, corrected q = 0.036), site 2 (monotherapy: 2.55 ± 0.43; multitherapy: 3.13 ± 0.77; *p* = 0.024, corrected q = 0.036), site 4 (monotherapy: 2.64 ± 0.65; multitherapy: 3.55 ± 1.29; *p* = 0.028, corrected q = 0.036) and in the average of all sites (monotherapy: 2.66 ± 0.37; multitherapy: 3.28 ± 0.81; *p* = 0.017, corrected q = 0.036) (Supplementary Figure 7A), while no differences were seen in *BDNF*-IV (Supplementary Figure 7B) nor in MDD patients (Supplementary Figure 7C, D).

### miRNA expression

#### miRNome

126 miRNAs have been identified to be in common between the CTRL, OCD, and MDD groups through a miRNome analysis. A fold-change scale is presented in Supplementary Figure 8 showing miRNAs abundance in OCD and MDD patients with respect to the CTRL values, considered to be 1. Overexpressed miRNAs are depicted in red, while the underexpressed are depicted in blue. Among the 126 miRNAs here, three miRNAs (*miR-16-5p*, *miR-29a-3p*, *miR-202-3p*) were upregulated, whereas *miR-191-5p* was downregulated in both OCD and MDD patients when compared to CTRLs.

#### miRNAs expression by qRT-PCR

We thus focused the attention on these 4 miRNAs, and we confirmed by *<u>qRT-PCR</u>* that *miR-16-5p* is statistically significantly upregulated in OCD when compared to CTRL group (OCD: 4.35 ± 4.02; CTRL: 1.19 ± 0.16; *p* = 0.002) (Figure 4A), but not in MDD (Figure 4E). We also confirm the overexpression of *miR-29a-3p* in saliva from both OCD (OCD: 2.47 ± 0.67; CTRL: 1.08 ± 0.09; *p* = 0.017) and MDD (MDD: 2.13 ± 0.36; CTRL: 1.08 ± 0.09; *p* = 0.014) subjects compared to CTRL group (Figure 4B, F). While no significant differences have been observed in *miR-191-5p* expression in OCD (Figure 4C), a significant downregulation was reported in MDD subjects when compared to CTRLs (MDD: 0.60 ± 0.10; CTRL: 1.18 ± 0.16; *p* < 0.001) (Figure 4G). *miR-202-3p* expression did not result altered in both OCD and MDD with respect to CTRLs (Figure 4D, H). Stratifying miRNA expression for TMS treatment no significant differences were observed in OCD subjects (Supplementary Figure 9A, B). Instead, there is an increase in *miR-29a-3p* expression following TMS in MDD group (CTRL: 1.08 ± 0.42; post-TMS MDD: 2.49 ± 1.67; adjusted *p* = 0.046) (Supplementary Figure 9C), and a significant decrease in *miR-191-5p* expression in both pre- and post-TMS MDD groups compared to CTRLs (CTRL: 1.18 ± 0.79; pre-TMS MDD: 0.61 ± 0.52; adjusted *p* = 0.026; post-TMS MDD: 0.59 ± 0.30; adjusted *p* = 0.035) (Supplementary Figure 9D). Stratifying data for the pharmacotherapy within OCD and MDD patients we didn’t report any significant differences (Supplementary Figure 10).

**Figure 4.**
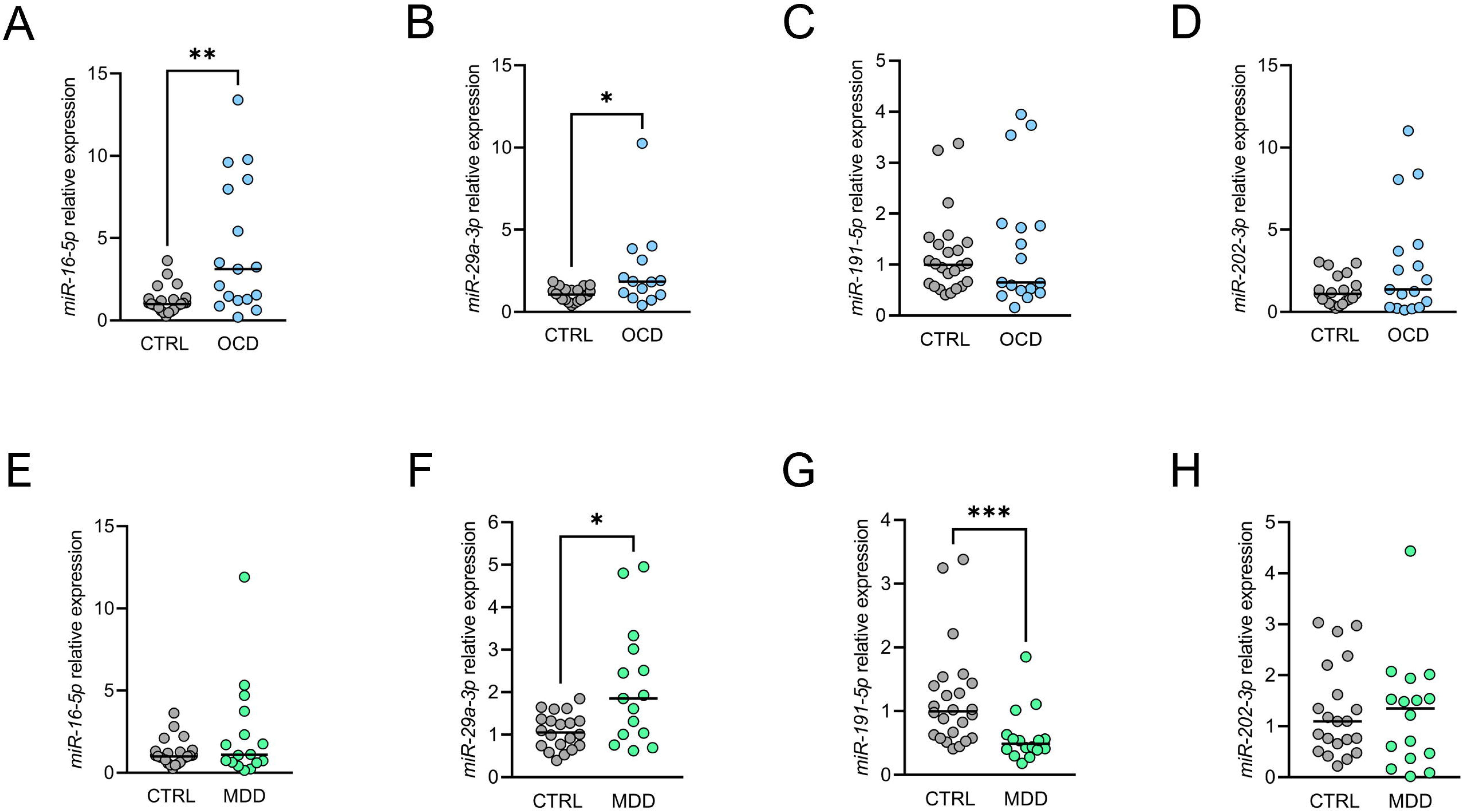
Exosomal miRNAs expression in saliva samples. *miR-16-5p* expression comparing OCD (**A**) and MDD (**E**) patients with CTRLs. *miR-29a-3p* expression comparing OCD (**B**) and MDD (**F**) patients with CTRLs. *miR-191-5p* expression comparing OCD (**C**) and MDD (**G**) patients with CTRLs. *miR-202-3p* expression comparing OCD (**D**) and MDD (**H**) patients with CTRLs. Data are presented as medians. (Mann-Whitney test, *p* < 0.05 considered to be significant; CTRL n=24; OCD n=17; MDD n=16).

### Correlation analysis

Phyla abundances correlated with methylation levels at *BDNF* gene (exon I and exon IV), miRNA levels and clinal parameters for both OCD and MDD (Figure 5). For details on correlation analysis at the genus level, please refer to Supplementary Figure 11A, B. Briefly, significant correlations were observed between Firmicutes abundances with *BDNF* methylation levels (inverse) and *miR-29a-3p* (direct) in OCD. Moreover, *BDNF* methylation at specific CpG sites inversely correlated with *miR-16-5p* and *miR-29a-3p* as well as illness duration (years from the onset) and age of onset. *miR-29a-3p* also inversely correlated with the Y-BOCS score. In MDD patients, an inverse correlation between Actinobacteria and Firmicutes with *BDNF* methylation and *miR-191-5p* was observed. Firmicutes abundance also directly correlated with the duration of illness (years from MDD onset). Bacteroidetes and Proteobacteria abundances directly correlated with *BDNF* methylation, and Proteobacteria also directly correlated with *miR-191-5p* expression. *BDNF* methylation also correlated directly with *miR-191-5p* levels, illness duration and the age of MDD onset.

**Figure 5.**
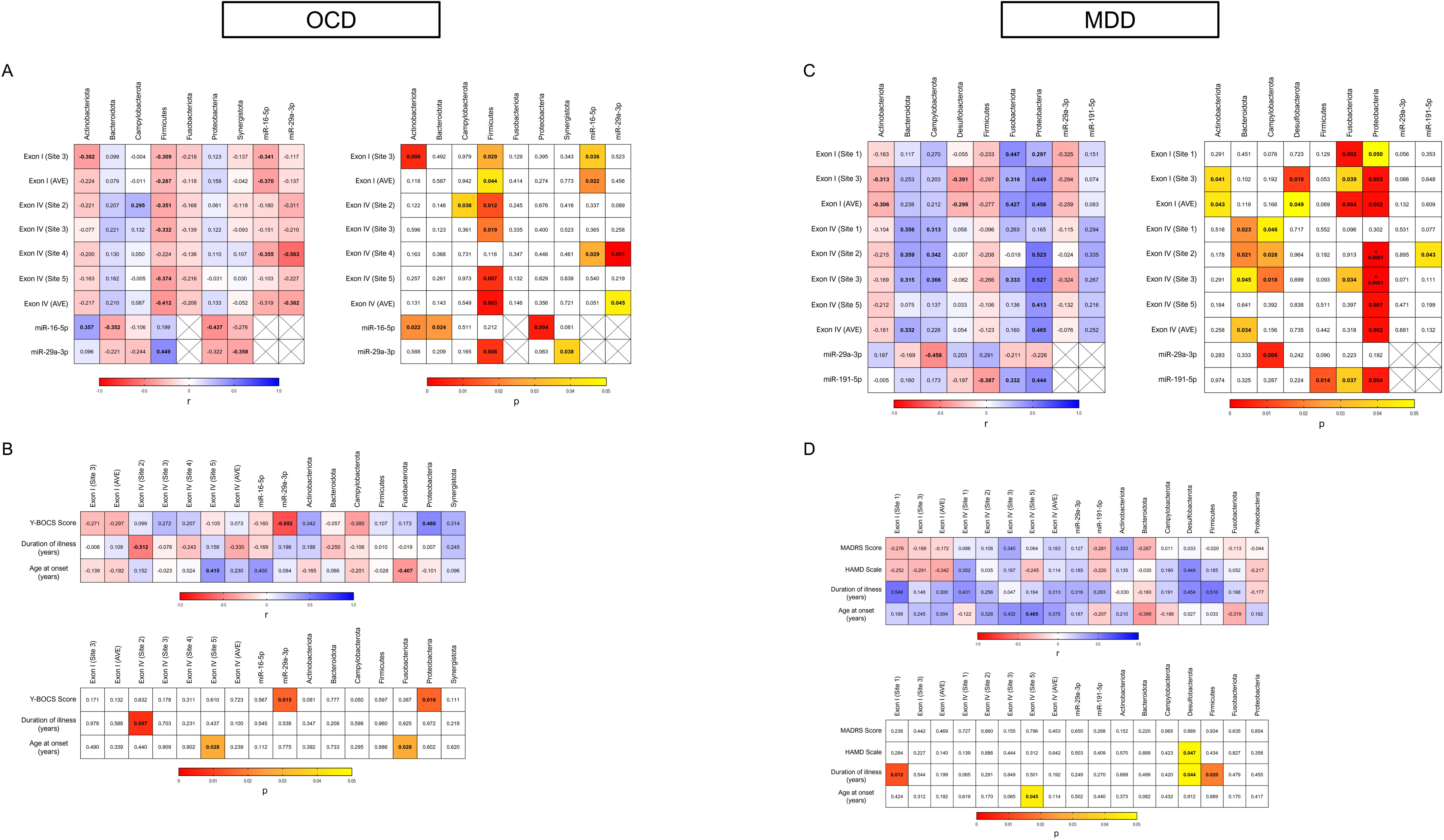
Correlation analysis between *BDNF* methylation levels, miRNAs, microbiota abundance (phyla) and clinical data in saliva from CTRL, OCD and MDD individuals. (**A, C**) Heat maps representing the correlation analysis between *BDNF* methylation levels, selectively altered miRNAs and phyla abundance in OCD and in MDD respectively. (**B, D**) Heat maps representing the correlation analysis of *BDNF* methylation levels, selectively altered miRNAs, phyla abundance with clinical data in OCD and in MDD respectively. Cells filled in blue to red gradient of the heat maps represent Spearman’s r; cells filled in yellow to red gradient represent *p* values (empty cells represent *p* values greater than 0.05). Statistically significant data are shown in bold.

## Discussion

The first main result of this study is that we show significant differences in salivary microbial diversity between OCD patients and healthy controls; in particular, a significant reduction in species evenness (alpha diversity: Simpson Index and Shannon Entropy), whereas no alpha diversity changes were seen in MDD when compared to controls. No significant differences in species richness (Chao1) in both OCD and MDD groups with respect to the CTRL group was observed, even if there is a tendency towards a reduction. Other studies previously analyzed alpha diversity in MDD, however, many of these did not show any alterations [81]. Our findings in OCD are unprecedented. Notably, lower alpha diversity has previously been reported in individuals with attention deficit hyperactivity disorder [82] and autism spectrum disorder (ASD) [83], both of which share etiological overlap with OCD [84].

In analyzing beta diversity, we report a significant variation in microbial communities in MDD when compared to controls, whereas no significant dissimilarity has been found in OCD subjects. A lower gut microbiome diversity was already observed in subjects self-reporting depression [85] as well as in diagnosed depression [86]. Moreover, changes in beta-diversity have been also reported based on the severity of MDD [87] and shown to be associated with depressive symptoms in different ethnic groups [88].

Analyzing differential abundance in microbial taxa in more depth, we show that 7 phyla (Actinobacteriota, Bacteroidota, Campylobacterota, Firmicutes, Fusobacteriota, Proteobacteria, Synergistota) and 11 bacterial genera (*Rothia*, *Alloprevotella*, *Campylobacter*, *Streptococcus*, *Gemella*, *Catonella*, *W5053*, *Sneathia*, *Simonsiella*, *Enhydrobacter*, *Pyramidobacter*) were significantly altered in OCD patients when compared to controls. Actinobacteriota, Firmicutes, Proteobacteria and Synergistota phyla were more abundant in OCD patients, while Bacteroidota and Campylobacterota were found to have reduced abundance. Of relevance, higher levels of Actinobacteriota and Firmicutes have already been reported in previous studies in OCD [25, 28] with *Streptococcus* resulted in being the most abundant bacteria within Firmicutes.

Considering MDD patients when compared to controls, we observed alterations in 7 phyla (Actinobacteriota, Bacteroidota, Campylobacterota, Desulfobacterota, Firmicutes, Fusobacteriota, Proteobacteria) and 38 genera, among which 19 (*Gardnerella*, *Scardovia*, *Rothia*, *Olsenella*, *Cryptobacterium*, *Slackia*, *Prevotellaceae UCG-004*, *Desulfobulbus*, *Asteroplasma*, *Lacticaseibacillus*, *Lactobacillus*, *Limosilactobacillus*, *Pseudoramibacter*, *Howardella*, *Shuttleworthia*, *Parvimonas*, *Peptoniphilus*, *Selenomonas*, *Dialister*) were more abundant and 19 (*Pseudopropionibacterium*, *F0058*, *Alloprevotella*, *Bergeyella*, *Campylobacter*, *Catonella*, *Jonhsonella*, *Ruminococcus*, *Peptococcus*, *Eubacterium yurii*, *Oceanvirga*, *Lautropia*, *Alysiella*, *Kingella*, *Neisseria*, *Simonsiella*, *Actinobacillus*, *Aggregatibacter*, *Haemophilus*) reduced in MDD compared to controls. It was previously reported that the proportion of Bacteroides, Proteus and Actinomycetes was significantly higher in MDD [89], whereas was observed an increase in Actinobacteriota and Firmicutes and a decrease in Bacteroidota and Proteobacteria in gut microbiota from MDD patients [90]. Consistent with what was reported by Chung et al., we show a significant increase in Actinobacteriota and Firmicutes, and a decrease in Bacteroidota and Proteobacteria abundances.

Although a different salivary microbiota composition has been observed in both OCD and MDD patients, no significant differences were seen when stratifying both clinical groups for TMS treatment. The only exceptions could be represented by Firmicutes where, in the post-TMS group, two genera *Gemella* and *Streptococcus* seemed to be more abundant in OCD patients but not in pre-TMS group, and one genus (*Catonella)* was significantly altered in the pre-TMS MDD group, which returned to control level after the treatment in the post-TMS group. However, data should be confirmed with a larger sample size study.

These results point to a possible oral microbiota signature in MDD and OCD. To better clarify how the dynamic community of microorganisms coexist in symbiosis with the host and microbiome-derived metabolites, regulating the host genome, it is of relevance to analyze the role of epigenetic mechanisms.

We focused our attention on the transcriptional regulation of *BDNF*, due to the relevance of microbiota in neurogenesis and the production of neurotrophins [91], since its epigenetic regulation has been reported to be involved in various psychiatric disorders, including OCD and MDD [92]. We report significant reductions in *BDNF* DNA methylation in both OCD and MDD groups when compared with controls, for CpG sites in two regulatory regions located upstream Exon I and Exon IV (Figure 3). This is consistent with previous findings, where increased levels of protein plasma BDNF levels have been reported in patients with OCD when compared to controls [93, 94]; however, other studies have reported no differences [95] or lower levels [37, 96]. Moreover, decreased levels in *BDNF* gene DNA methylation and increased hydroxymethylation have been reported in saliva samples of OCD patients [34]. Several studies have reported reductions in BDNF levels in plasma/ serum of patients suffering from MDD [97, 98], and higher levels of *BDNF* promoter methylation associated with the reduced cortical thickness and also lower BDNF levels in peripheral blood in patients with MDD [92, 99, 100]. In contrast, we show a strong significant hypomethylation of the *BDNF* gene in saliva from patients diagnosed with MDD, suggesting higher levels of BDNF. However, previous studies also reported hypomethylation at a specific *BDNF* CpG site in patients with MDD compared with healthy controls [36]. Interestingly, TMS treatment appears to have a slight effect on the methylation pattern of *BDNF* in exon IV in both OCD and MDD patients. Further, we report a significantly higher methylation level at CpG site 2 in *BDNF* exon IV in MDD patients, demonstrating a positive response to the treatment, and thus suggesting a possible role for TMS in epigenetic modulation. However, more samples are needed to confirm these data.

Here we show that *miR-16-5p* was upregulated in OCD, *miR-29a-3p* was overexpressed in both OCD and MDD, and *miR-191-5p* was downregulated in MDD. miRNAs are known to reduce gene expression by multiple pathways and modalities and are self-assembling in effector complexes such as miRNP, miRgonaute, or miRISC binding at the mRNA target [101]. It has been also reported that some miRNAs can upregulate gene expression in specific cell types and conditions with distinct transcripts and proteins [102, 103]. However, we report here that TMS does not revert miRNAs expression pattern in both OCD and MDD patients.

We wanted to see if alterations in microbial abundance in OCD and MDD might be correlated to the epigenetic alterations in *BDNF* gene regulation. Of relevance in OCD, we observed an inverse correlation between firmicutes abundance and *BDNF* DNA methylation. Bacteria belonging to Firmicutes phyla are the major producers of butyrate [104], a short-chain fatty acid known to inhibit histone deacetylases (HDACs), and histone acetylation closely interacts with DNA methylation inducing demethylation [105]. On the other hand, analyzing MDD data, it can be observed that Fusobacteria and Proteobacteria known to have a key role in DNA methylation [106, 107], are significantly directly correlated with *BDNF* DNA methylation and this might relate to bacteria metabolites acting as methyl donors [108, 109]. Moreover, Firmicutes abundance in OCD and Fusobacteria, Proteobacteria and, again, Firmicutes levels in MDD are directly correlated with *miR-191-5p* expression, and thus again associated with *BDNF* gene regulation.

Recently studies are showing that microbiota can affect host miRNA expression [110] as well as miRNAs can regulate bacterial genes [111]. Nevertheless, the precise mechanisms remain unclear.

In conclusion, the present study shows a unique salivary microbiome signature in individuals with OCD and MDD and suggests a microbiota-host epigenetic axis. These microbial alterations seem to relate to specific changes in the transcriptional regulation of *BDNF*. Our hypothesis is thus that it is possible to identify specific biomarkers for MDD and OCD using saliva as a non-invasive collection method. to obtain genomic material. This approach also represents a good strategy to easily monitor different samples with different ages (children, elderly) and/or sensitive subjects during the development of the disease. These methodological and mechanistic insights serve as a foundation for future research and provide avenues for the development of specific therapeutic interventions for these two prevalent psychiatric conditions targeting epigenetic regulation and microbiota analysis.

## Limitation of the study

This study has limitations that should be acknowledged. First, the small sample size of participants even if a power analysis was performed suggesting a power > 0.8 for the epigenetic analysis. We also observed potential effects of transcranial magnetic stimulation (TMS) on microbial and epigenetic markers. However, again, the small sample size of participants undergoing TMS may have influenced the significance and interpretation of these effects. A larger cohort is needed to validate these results and improve their robustness. Despite these limitations, our study highlights important connections between oral microbiota, epigenetic regulation, and psychiatric disorders, providing a foundation for future research into non-invasive biomarkers and targeted therapies.

## Data Availability

All data produced in the present study are available upon reasonable request to the authors.

## CRediT AUTHORSHIP CONTRIBUTION STATEMENT

A.G. and C.D.A., Conceptualization; A.G. and C.D.A., Writing - original draft; A.G., M.V., E.G., F.M., N.G., F.K., Investigation; A.G., M.V., K.J.O., E.G., F.K. Formal analysis and Data curation; C.D.A, L.S., J.F.C., B.D.O., Resources; M.V., K.J.O., E.G., F.M., M.P., V.G., L.S., J.F.C., B.D.O., and C.D.A. Supervision. We confirm that the manuscript has been read and approved by all named authors. We confirm that the order of authors listed in the manuscript has been approved by all named authors.

## AKNOWLEDGEMENT

This research was partially funded by European Union – Next Generation EU. Project Code: ECS00000041; Project CUP: C43C22000380007; Project Title: Innovation, digitalization and sustainability for the diffused economy in Central Italy – VITALITY; by Italian Minister of the University and Research, PRIN 2022 Project code: 2022K7YKTY to C.D. This work was supported, in part, by research grants from Science Foundation lreland (SFI) to APC Microbiome lreland through the Irish Government’s National Development Plan SFI/12/RC/2273_P2.

## CONFLICT OF INTEREST

We wish to confirm that there are no known conflicts of interest associated with this publication and there has been no significant financial support for this work that could have influenced its outcome.

We confirm that we have given due consideration to the protection of intellectual property associated with this work and that there are no impediments to publication, including the timing of publication, with respect to intellectual property. In so doing we confirm that we have followed the regulations of our institutions concerning intellectual property.

We hereby certify:

- that the paper is original and result of our research in full, does not contain earlier published sections, and as such does not infringe upon anyone’s copyright nor violate any proprietary rights;
- that we take full responsibility for the conducted research, data analysis and interpretation, and conclusion;
- the used bibliographical references are clearly stated in the paper itself and in the list of references;
- this paper (or any section thereof) has not been published and will not be sent for publishing to another journal or publication prior to receiving notification whether the paper will be published in Molecular Psychiatry.

