## Supplementary material for "DISTINCT SALIVARY MICROBIOTA PROFILES, *BDNF* DNA METHYLATION AND *MIR-16-5P*, *MIR-29A-3P*, *MIR-191-5P* ALTERATIONS IN OBSESSIVE-COMPULSIVE DISORDER AND MAJOR DEPRESSIVE DISORDER": Supplementary material_MOLPSY2025.docx

**Microbiota-Host Epigenetic Axis In Saliva**

Antonio Girella^1^, MSc; Matteo Vismara^2^, MD; Kenneth J. O'Riordan^3^, PhD; Eoin Gunnigle^3^, PhD; Francesca Mercante^1^, MSc; Nicolaja Girone^2^, MD; Mariangela Pucci^1,4^, PhD; Valentina Gatta^5,6^, PhD; Fani Konstantinidou^5,6^, PhD; Liborio Stuppia^5,6^, PhD; John F. Cryan^3,7^, PhD; Bernardo Dell’Osso^2,8^, MD; Claudio D’Addario^1,9,#^, PhD

^1^ Department of Bioscience and Technology for Food, Agriculture and Environment, University of Teramo, Teramo, 64100, Italy

^2^ Department of Psychiatry, Department of Biomedical and Clinical Sciences “Luigi Sacco”, University of Milan, ASST Fatebenefratelli-Sacco, Milan, 20019, Italy

^3^ APC Microbiome Ireland, University College Cork, Cork, Ireland

^4^ Department of Biosciences and Nutrition, Karolinska Institute, Huddinge, Sweden

^5^ Department of Psychological Health and Territorial Science, School of Medicine and Health Sciences, “G. D’Annunzio”, University of Chieti-Pescara, Chieti, 66100, Italy

^6^ Unit of Molecular Genetics, Center of Advanced Studies and Technology (CAST), “G. D’Annunzio”, University of Chieti-Pescara, Chieti, 66100, Italy

^7^ Department of Anatomy and Neuroscience, University College Cork, Cork, Ireland

^8^ “Aldo Ravelli” Center for Nanotechnology and Neurostimulation, University of Milan, Milan, Italy

^9^ Department of Clinical Neuroscience, Karolinska Institute, Stockholm, 10316, Sweden

\# Corresponding author:

Claudio D'Addario,

Department of Bioscience and Technology for Food, Agriculture and Environment, University of Teramo, Via Renato Balzarini, 1, 64100 Teramo, Italy.

Phone number: +39 0861 266877

**Supplementary information**

### **Materials and Methods**

#### **TMS stimulation**

Exclusion criteria to rTMS involved the presence of neurological disorders (epilepsy or familiarity for epilepsy, previous significant head trauma, brain surgery, loss of consciousness for minimum 15 minutes), pregnancy or lactation, significant medical and/or psychiatric comorbidities, substance abuse in the last 3 months, pacemakers or electrical stimulation devices, metallic clips, severe cardiac disorders, hypertension and sleep apnea. Stimulation was administered using a MagVenture Magpro R30 high-performance TMS stimulator connected to a figure-of-eight coil (MagVenture Company, Farum, DK). The identification of the resting motor threshold (RMT) and of the target area was performed according to standard procedures [1]. RMT was considered as the minimum TMS intensity producing a motor evoked potential in the contralateral abductor pollicis brevis in at least 50% of trials [2]. The target area was identified using the International 10-20 EEG system, with stimulation parameters chosen within those recommended by recent International Safety Guidelines [3]: for OCD left orbitofrontal cortex (OFC), 1 Hz frequency, intensity of stimulus 80% of RMT, 600 stimuli per session, 15 sessions of rTMS (5 sessions per week for 3 consecutive weeks); for MDD, left dorsolateral prefrontal cortex (DLPFC), 10 Hz frequency, intensity of stimulus 120% of RMT, 11 trains of 30 seconds each, interspersed by 30 seconds of pause (3000 stimuli per session), total 20 sessions of rTMS (2 sessions per day for 2 consecutive weeks).

#### **Supplementary Tables**

| ***Psychometric variables*** | **OCD (n=10)** | | |
| --- | --- | --- | --- |
|  | **Pre-TMS** | **Post-TMS** | **P value** |
| Y-BOCS score at assesment (mean ± SD) | 25.0 ± 8.42 | 18.75 ± 5.17 | 0.072 (parametric t-test) |
|  | **MDD (n=16)** | | |
| HAMD score at assesment (mean ± SD) | 17.7 ± 4.71 | 12.1 ± 5.22 | 0.022 (parametric t-test) |
| MADRS score at assesment (mean ± SD) | 24.6 ± 5.54 | 19.8 ± 8.04 | 0.137 (parametric t-test) |

**Supplementary Table 1.** Psychometric variables assessed before and after repetitive transcranial magnetic stimulation (TMS) in Obsessive-Compulsive Disorder (OCD, n=10) and Major Depressive Disorder (MDD, n=16) patients. Scores are presented as mean ± standard deviation. Parametric Welch’s test with *p* < 0.05 considered to be significant.

| ***BDNF* promoter** | **Primers** | **Sequence to analyze** | **Human (GRCh38:** **CM000673.2)** |
| --- | --- | --- | --- |
| **Exon I** | Included in the assay Hs_BDNF_08_PM PyroMark CpG Assay  (PM00155540) | t**CGCG**ctt**CG**a**CG**catgcag | Chromosome 11: 27722318-27722299 |
| **Exon IV** | F:  AGGTAGGGAGATTTTATGTTAGT  R-biotin: ACCCTAAAACCAAACTCTTCTAATAAAAAA  S:  AATGGGAAAGTGGGT | actcccacccactttcccattcac**CGCG**gagagggctgctct**CG**ctgc**CG**ctccccc**CG**g**CG**a | Chromosome 11: 27701639-27701577 |

**Supplementary Table 2.** Details of sequences, primers, and assays employed during the analysis of DNA methylation. F = forward primer; R-biotin = biotinylated reverse primer; S = sequencing primer.; Bold text = CpG sites analyzed.

| **Target miRNAs** | **miRCURY LNA miRNA Probe PCR Assay** | **miRBase Accession** | **Mature miRNA sequence** | **Annealing T (°C)** | **PCR product melting T (°C)** | **PCR efficiency (%)** |
| --- | --- | --- | --- | --- | --- | --- |
| *hsa-miR-16-5p* | YP00205702 | MIMAT0000069 | UAGCAGCACGUAAAUAUUGGCG | 58 | 73.2 ± 0.7 | 91.8 ± 2.6 |
| *hsa-miR-29a-3p* | YP00204698 | MIMAT0000086 | UAGCACCAUCUGAAAUCGGUUA | 58 | 72.4 ± 0.3 | 90.7 ± 1.8 |
| *hsa-miR-191-5p* | YP00204306 | MIMAT0000440 | CAACGGAAUCCCAAAAGCAGCUG | 58 | 73.3 ± 0.9 | 94.5 ± 0.7 |
| *hsa-miR-202-3p* | YP00205990 | MIMAT0002811 | AGAGGUAUAGGGCAUGGGAA | 58 | 73.1 ± 0.5 | 88.3 ± 3.4 |
| **References** |  |  |  |  |  |  |
| *U6* | YP02119464 | U6snRNA | Not provided from Qiagen | 58 | 72.3 ± 0.4 | 95.1 ± 1.4 |
| *U48* | YP00203903 | SNORD48 | Not provided from Qiagen | 58 | 75.4 ± 0.6 | 92.2 ± 1.6 |

**Supplementary Table 3.** Information about primers used for miRNAs qRT-PCR. Test annealing temperature (°C), PCR products melting temperature (°C) as well as PCR efficiency (%) for every test are reported.

### **Supplementary figures**

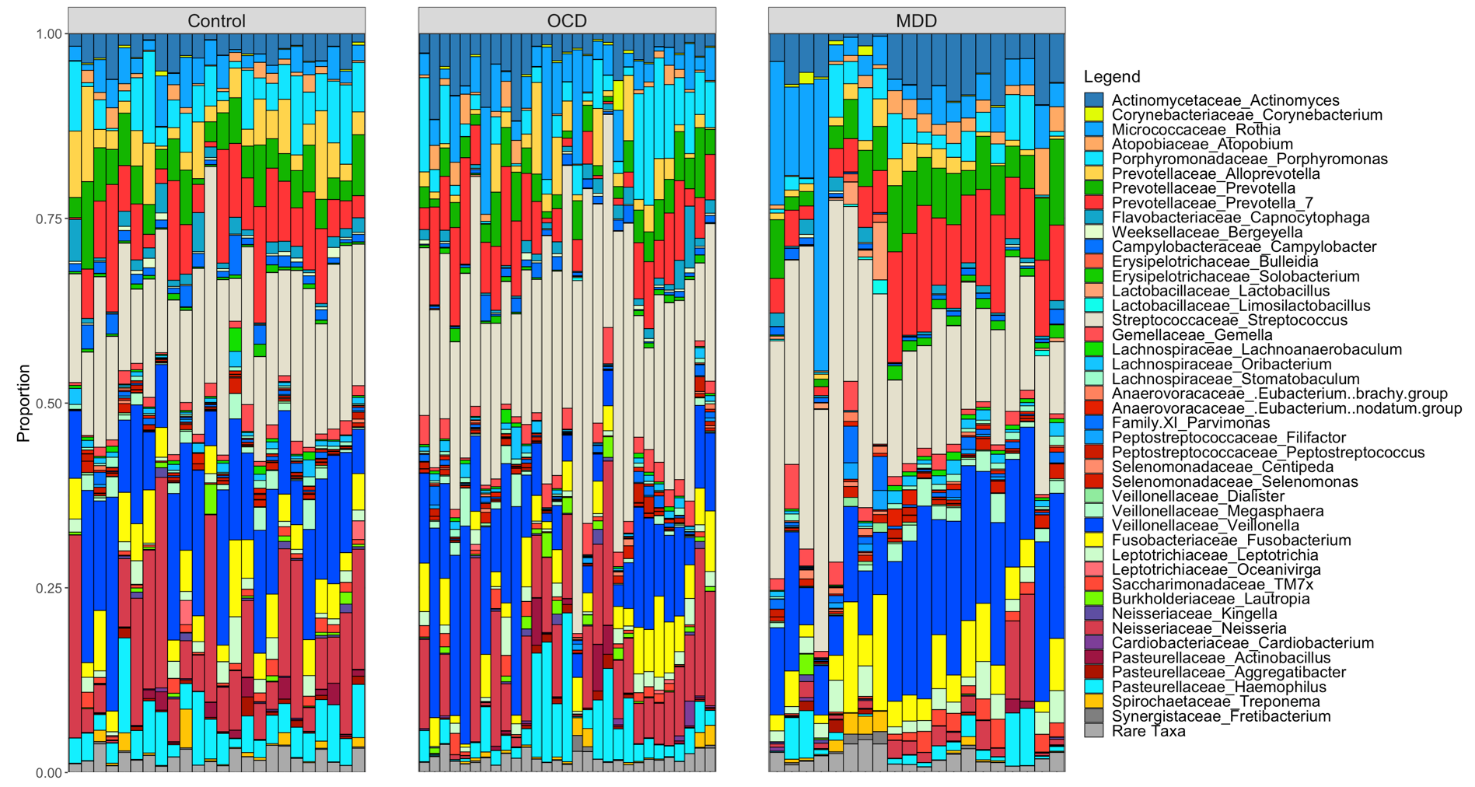

**Supplementary Figure 1.** Salivary microbiota composition. Mean relative abundance of major taxa in saliva from CTRL healthy individuals (n=24), OCD (n=29) and MDD patients (n=20). For relative abundance (%) please refer to “Figure S1 excel spreadsheet”.

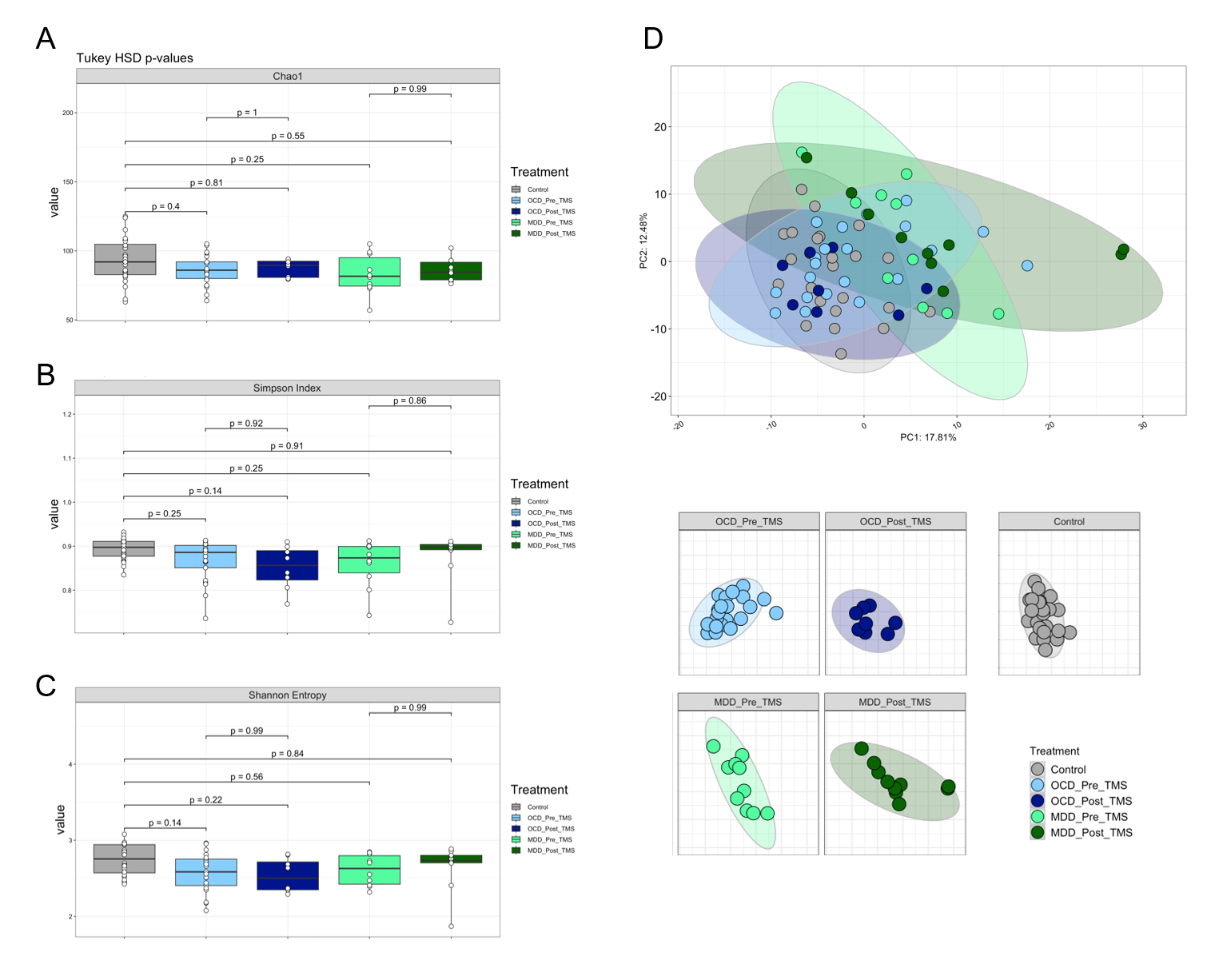

**Supplementary Figure 2.** Salivary microbiota composition in CTRL healthy individuals (n=24), OCD and MDD patients stratified for pre- and post-TMS treatment (OCD pre-TMS n=21; OCD post-TMS n=8; MDD pre-TMS n=10; MDD post-TMS n=10). Alpha diversity indices: (**A**) Chao1, (**B**) Simpson index and (**C**) Shannon entropy. (**D**) PCA plot of beta diversity of salivary microbiota of CTRLs, OCD and MDD patients stratified for TMS treatment. Data considered to be significant with a *p* value < 0.05 (Tukey-Honestly Significant Difference (HSD)).

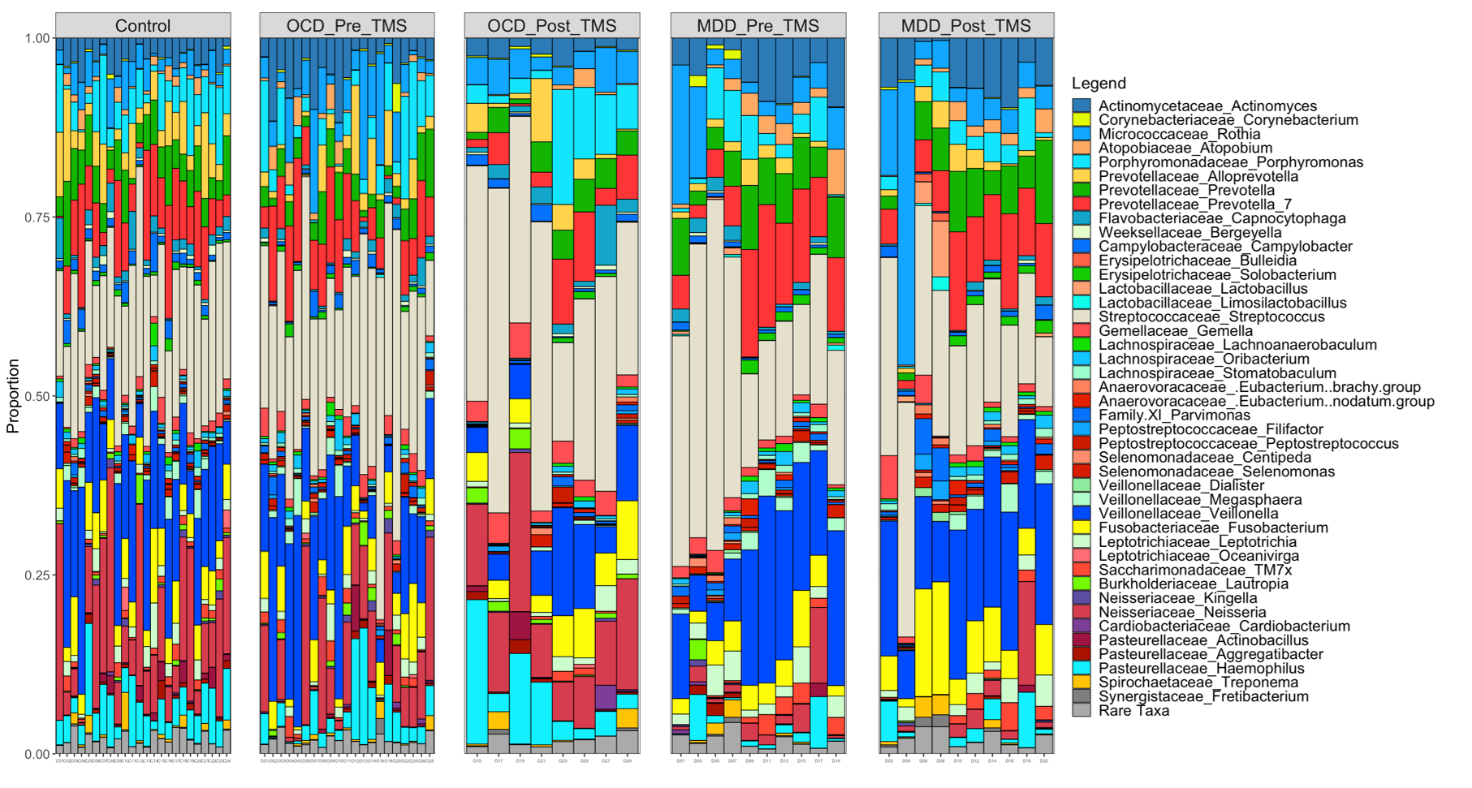

**Supplementary Figure 3.** Salivary microbiota composition. Mean relative abundance of major taxa in saliva from CTRL healthy individuals (n=24), OCD and MDD patients stratified for pre- and post-TMS treatment (OCD pre-TMS n=21; OCD post-TMS n=8; MDD pre-TMS n=10; MDD post-TMS n=10). For relative abundance (%) please refer to “Figure S3 excel spreadsheet”.

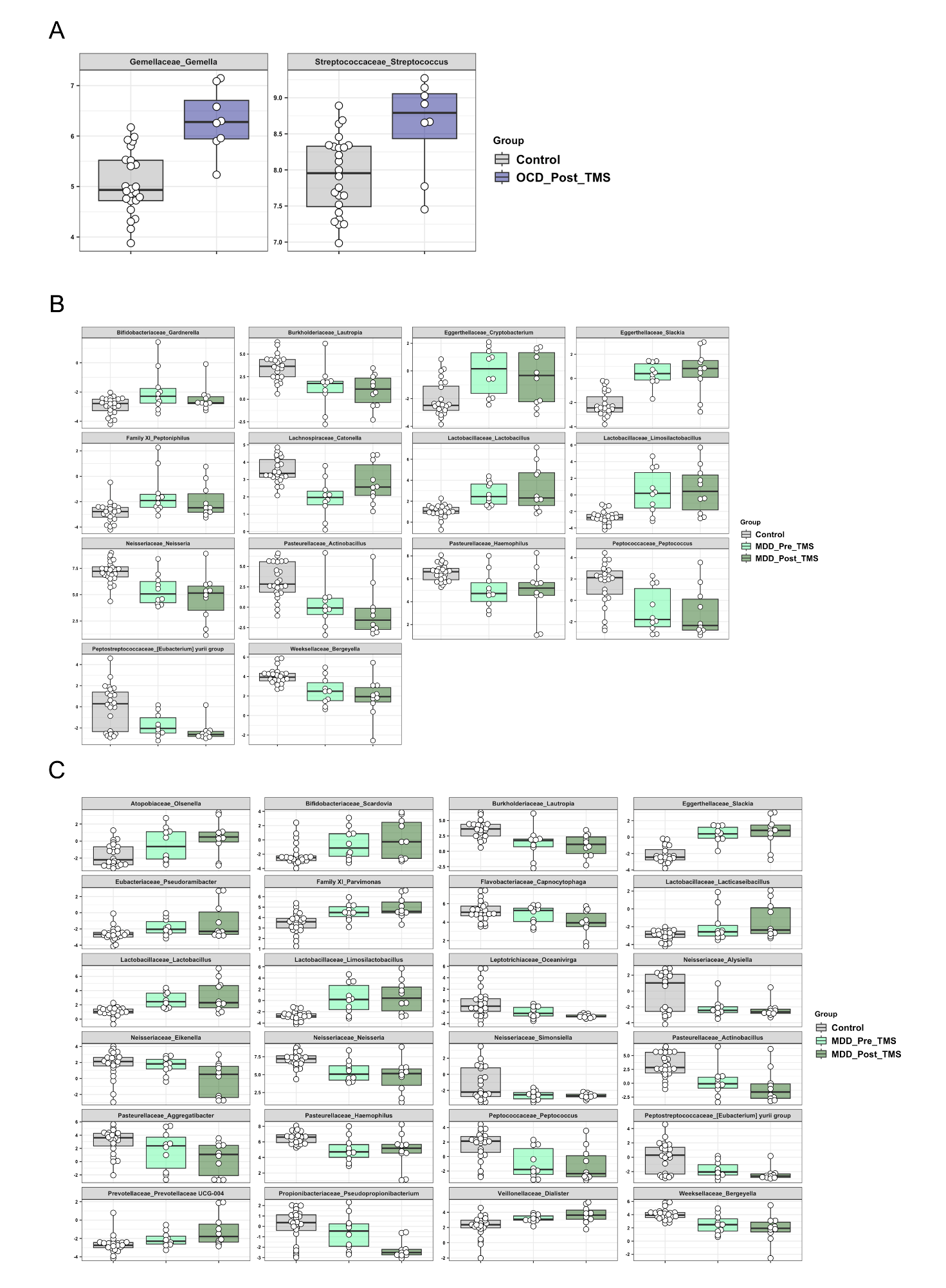

**Supplementary Figure 4.** Salivary microbiota composition in CTRL healthy individuals (n=24), OCD and MDD patients stratified for pre- and post-TMS treatment (OCD pre-TMS n=21; OCD post-TMS n=8; MDD pre-TMS n=10; MDD post-TMS n=10). (**A**) Differentially abundant genera from saliva of CTRL vs post-TMS OCD individuals. (**B**) Differentially abundant genera from saliva of CTRL vs pre-TMS MDD individuals. (**C**) Differentially abundant genera from saliva of CTRL vs post-TMS MDD individuals. Data considered to be significant with an adjusted p value < 0.05 (Tuckey-Generalised Linear Model (GLM) 10% FDR correction).

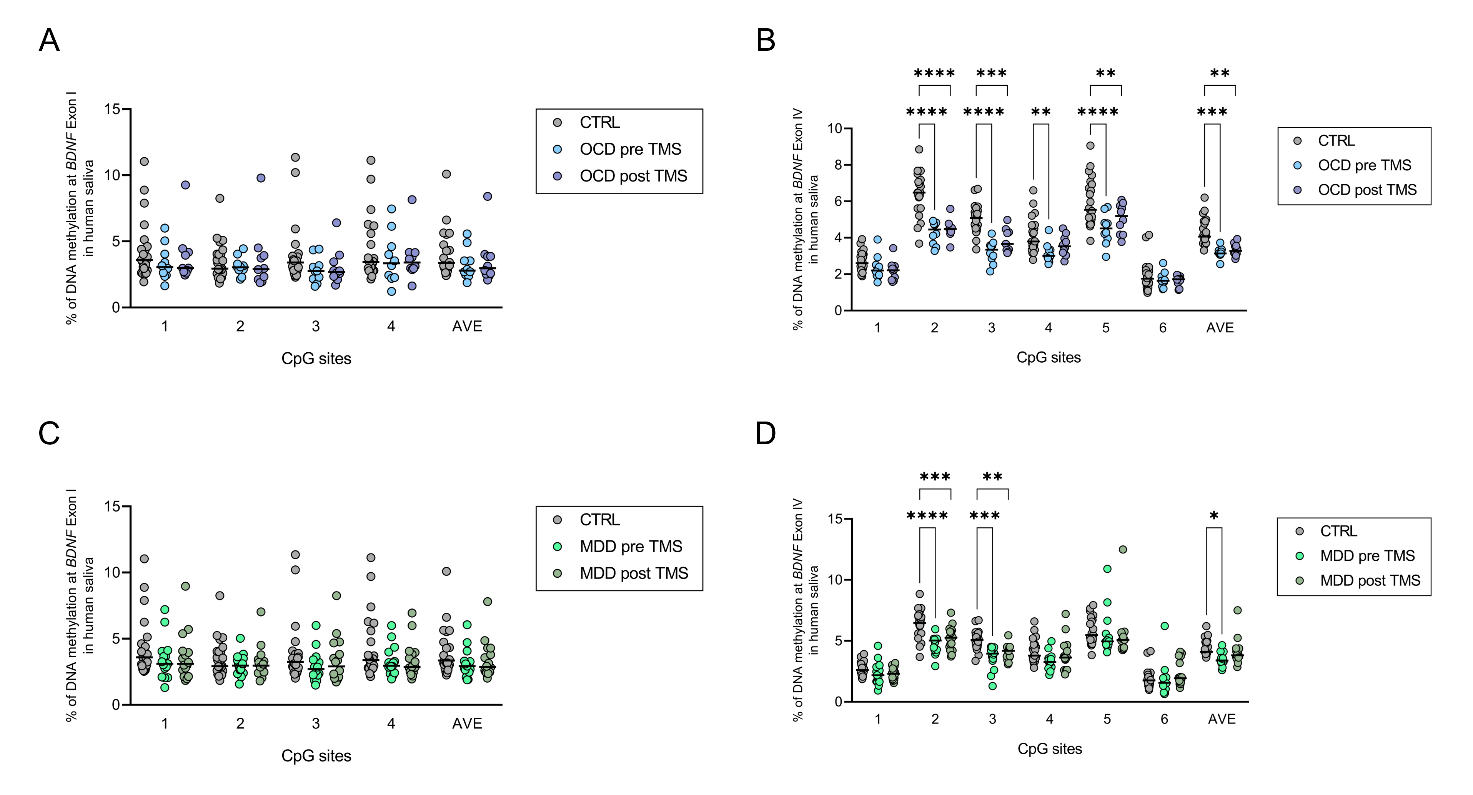

**Supplementary Figure 5.** BDNF gene regulation in saliva samples. (**A**) % of DNA methylation at BDNF exon I promoter (4 CpG sites) considering CTRLs and OCD patients underwent TMS treatment. (**B**) % of DNA methylation at BDNF exon IV promoter (6 CpG sites) considering CTRLs and OCD patients underwent TMS treatment (CTRL n=24; OCD pre-TMS n=10; OCD post-TMS n=10). (**C**) % of DNA methylation at BDNF exon I promoter (4 CpG sites) considering CTRLs and MDD patients underwent TMS treatment. (**D**) % of DNA methylation at BDNF exon IV promoter (6 CpG sites) considering CTRLs and MDD patients underwent TMS treatment (CTRL n=24; MDD pre-TMS n=16; MDD post-TMS n=16). Two-way ANOVA and FDR 10% BH correction, q < 0.1 considered to be significant. Data are represented as scatter dots with medians.

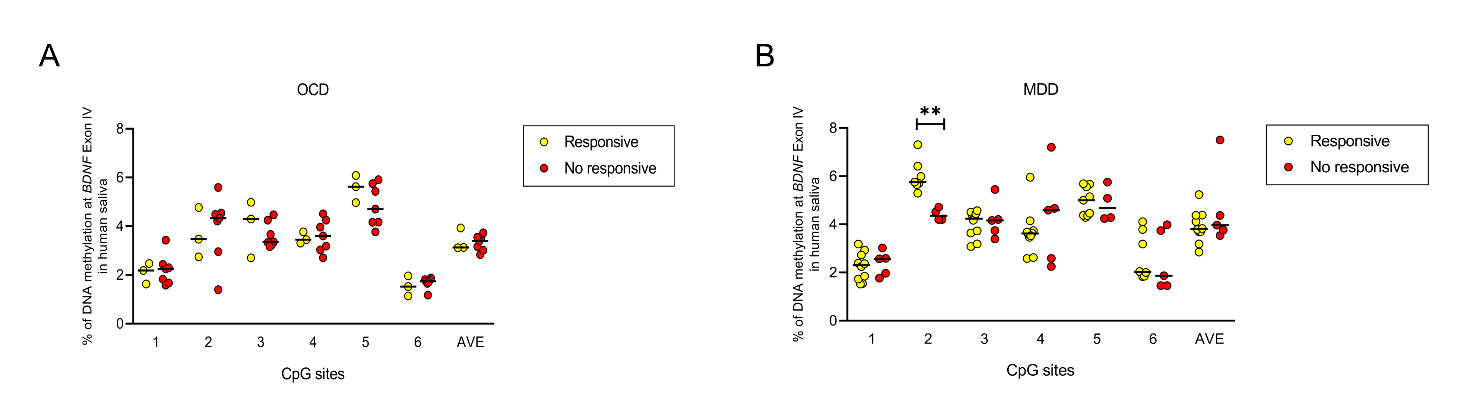

**Supplementary Figure 6.** BDNF gene regulation at Exon IV in saliva samples. (**A**) % of DNA methylation at BDNF exon IV promoter (6 CpG sites) considering post-TMS OCD responsiveness (responsive n=3; no responsive n=7). (**B**) % of DNA methylation at BDNF exon IV promoter (6 CpG sites) considering post-TMS MDD responsiveness (responsive n=10, no responsive n=6). Unpaired t-test and FDR 10% BH correction, q < 0.1 considered to be significant. Data are represented as scatter dots with medians.

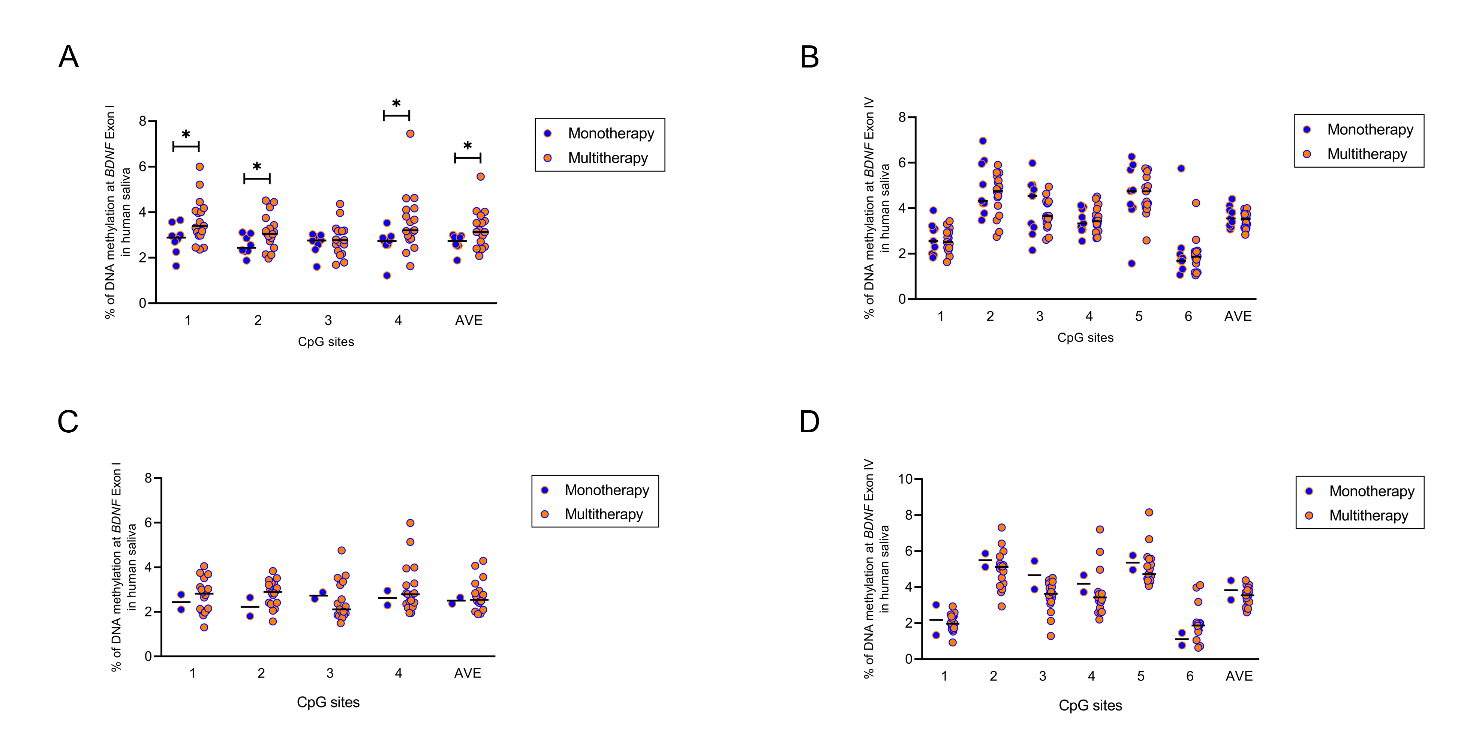

**Supplementary Figure 7.** BDNF gene regulation in saliva samples. (**A**) % of DNA methylation at BDNF exon I promoter (4 CpG sites) considering OCD patients stratified for pharmacological therapy. (**B**) % of DNA methylation at BDNF exon IV promoter (6 CpG sites) considering OCD patients stratified for pharmacological therapy (monotherapy n=7; multitherapy n=16). (**C**) % of DNA methylation at BDNF exon I promoter (4 CpG sites) considering MDD patients stratified for pharmacological therapy. (**D**) % of DNA methylation at BDNF exon IV promoter (6 CpG sites) considering MDD patients stratified for pharmacological therapy (monotherapy n=2; multitherapy n=18). Unpaired t-test and FDR 10% BH correction, q < 0.1 considered to be significant. Data are represented as scatter dots with medians.

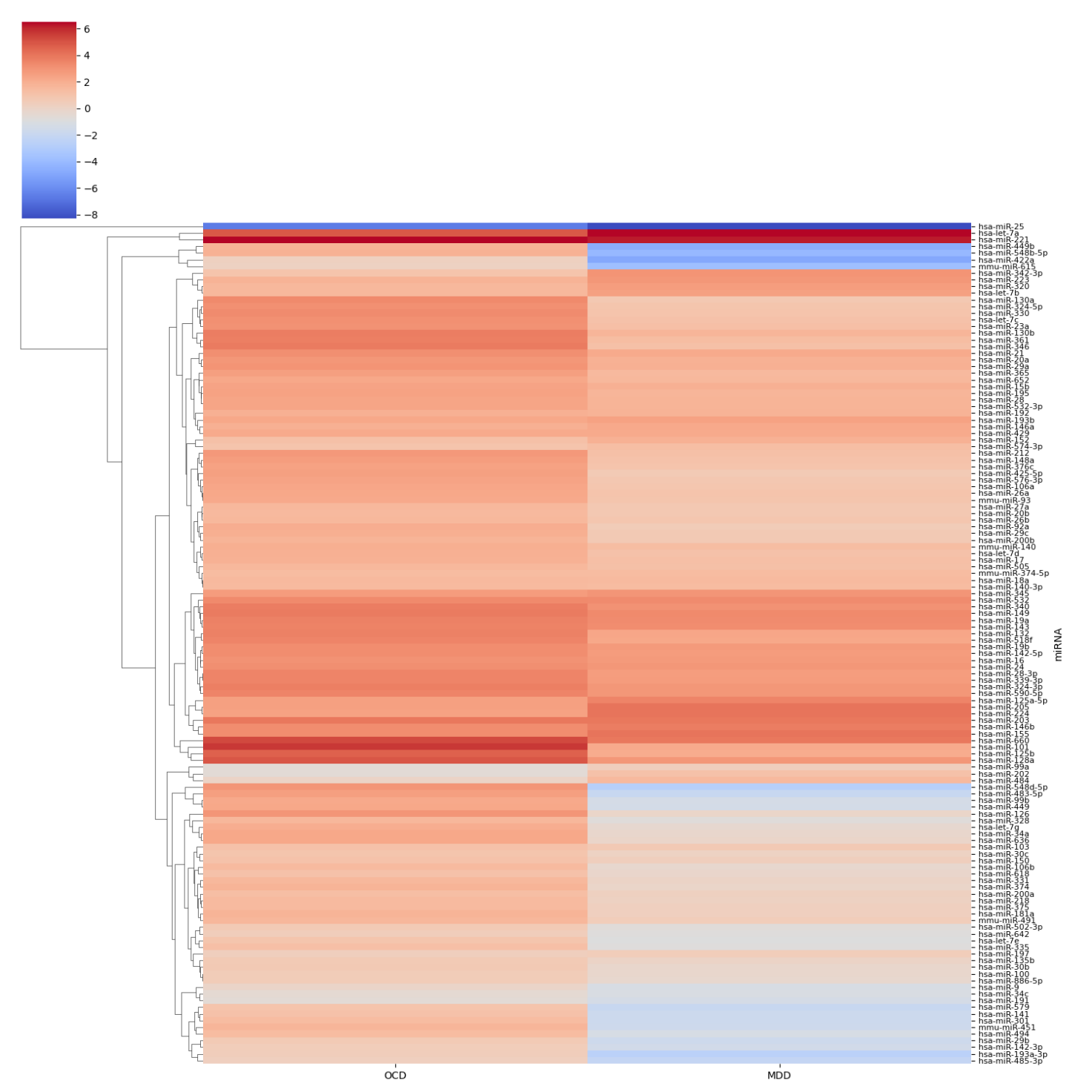

**Supplementary Figure 8.** miRNome cluster panel showing 126 miRNAs differentially regulated in saliva from OCD and MDD patients compared with CTRLs. A fold-change scale is reported: overexpressed miRNAs are depicted in red, while underexpressed ones are depicted in blue (CTRL’s miRNAs expression considered to be 1). Differences between groups were calculated by the Delta-Delta Ct (ΔΔCt) method and converted to fold change expressed as 2^(−ΔΔCt)^. For the fold-change differences please refer to “Figure S8 excel spreadsheet”.

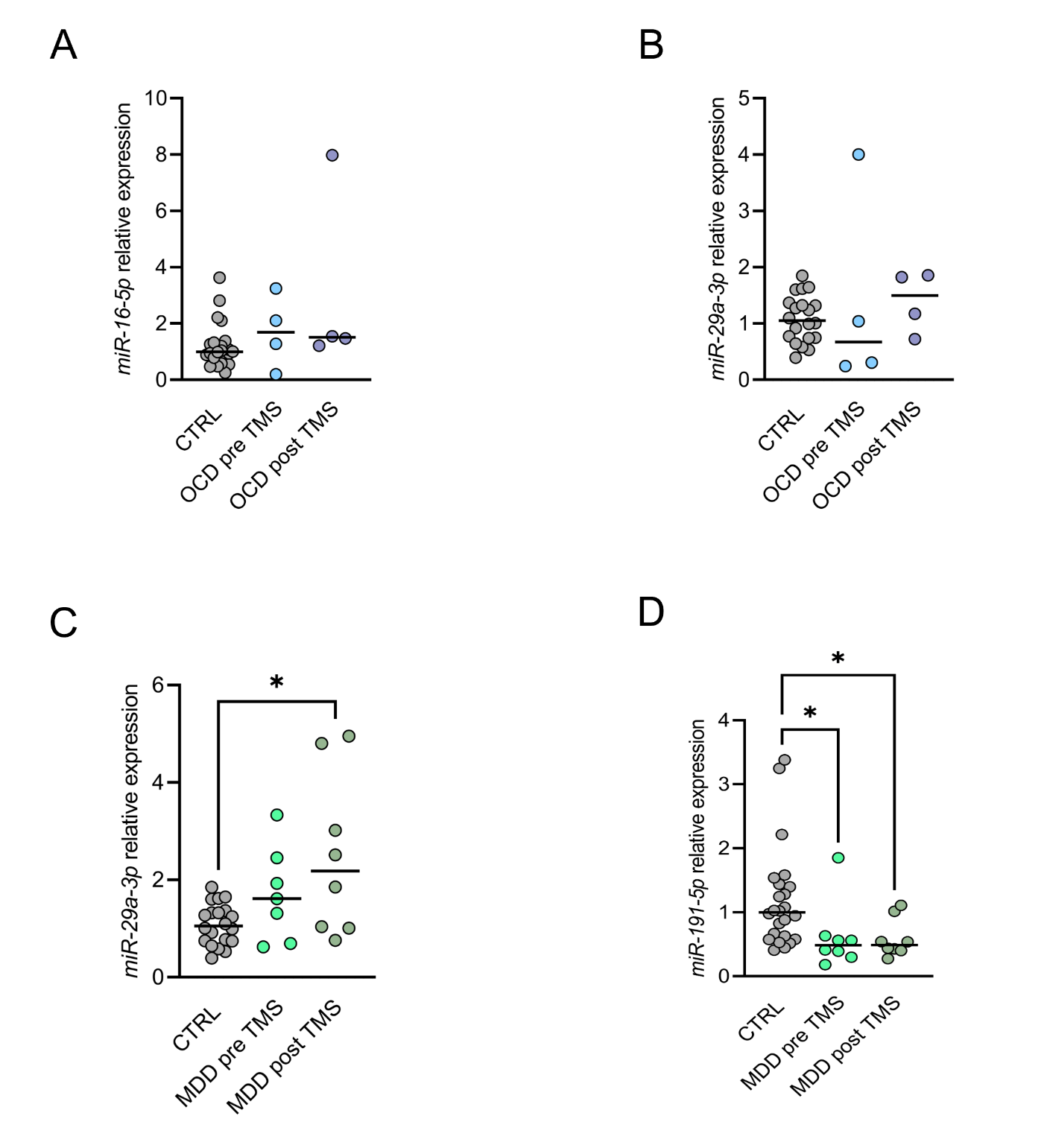

**Supplementary Figure 9.** Exosomal miRNAs expression in saliva samples. (**A**) miR-16-5p and (**B**) miR-29a-3p expression considering CTRLs (n=24) and OCD patients underwent TMS treatment (OCD pre-TMS n=4; OCD post-TMS n=4). (**C**) miR-29a-3p and (**D**) miR-191-5p expression considering CTRLs (n=24) and MDD patients underwent TMS treatment (MDD pre-TMS n=8; MDD post-TMS n=8). Kruskal-Wallis test with Dunn’s correction, p < 0.05 considered to be significant. Data are presented as dots with medians.

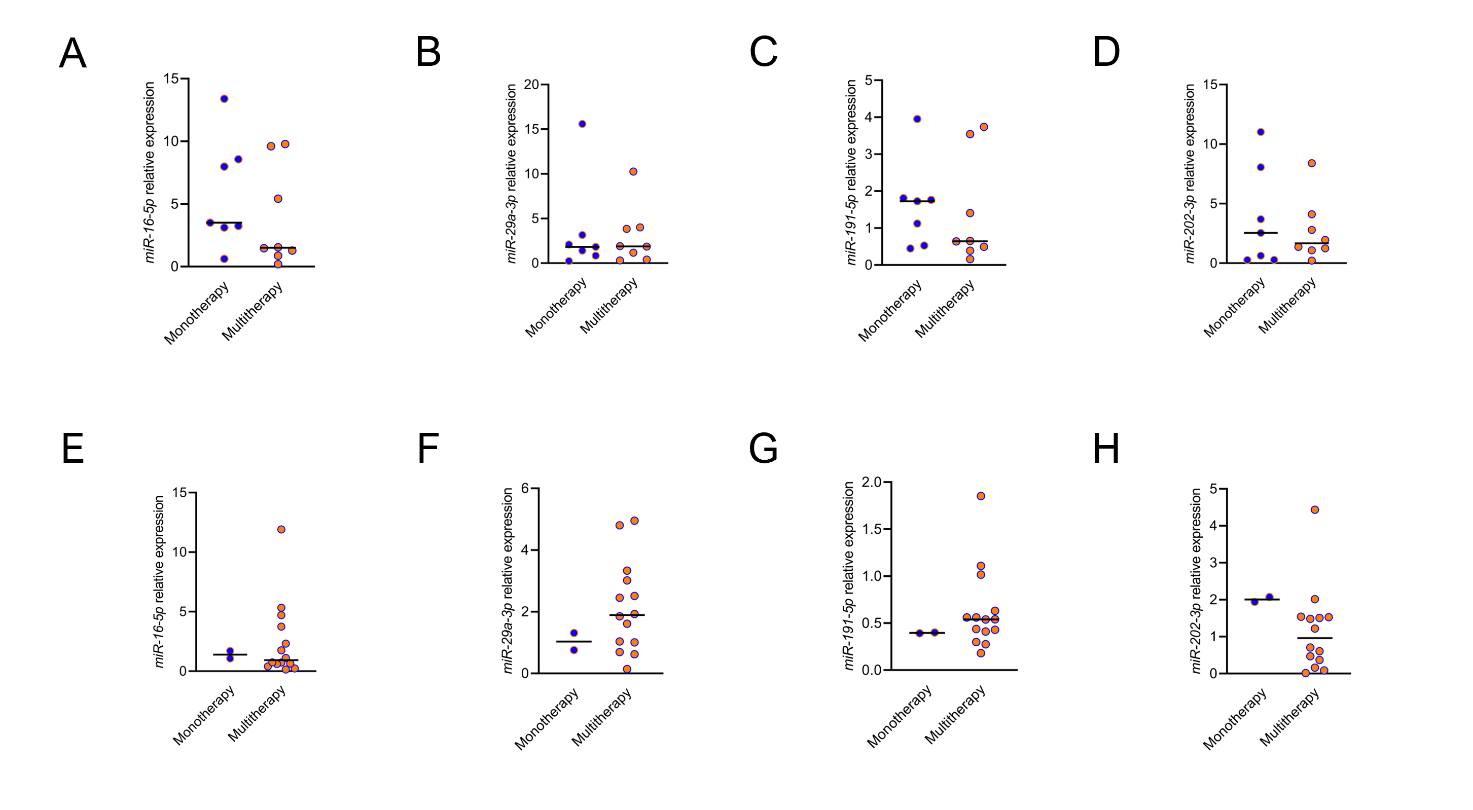

**Supplementary Figure 10.** Exosomal miRNAs expression in saliva samples. *miR-16-5p*, *miR-29a-3p*, *miR-191-5p* and *miR-202-3p* expression in (**A-D**) OCD patients and (**E-H**) MDD patients stratified for pharmacological therapy (OCD: monotherapy n=7; multitherapy n=8) (MDD: monotherapy n=2; multitherapy n=14). Mann-Whitney test, *p* < 0.05 considered to be significant. Data are presented as medians.

#### **Correlation analysis**

Identified microbiota targets were examined for their correlations with methylation levels at *BDNF* gene (exon I and exon IV), with significantly altered miRNAs and with clinal parameters (Supplementary Figure 6).

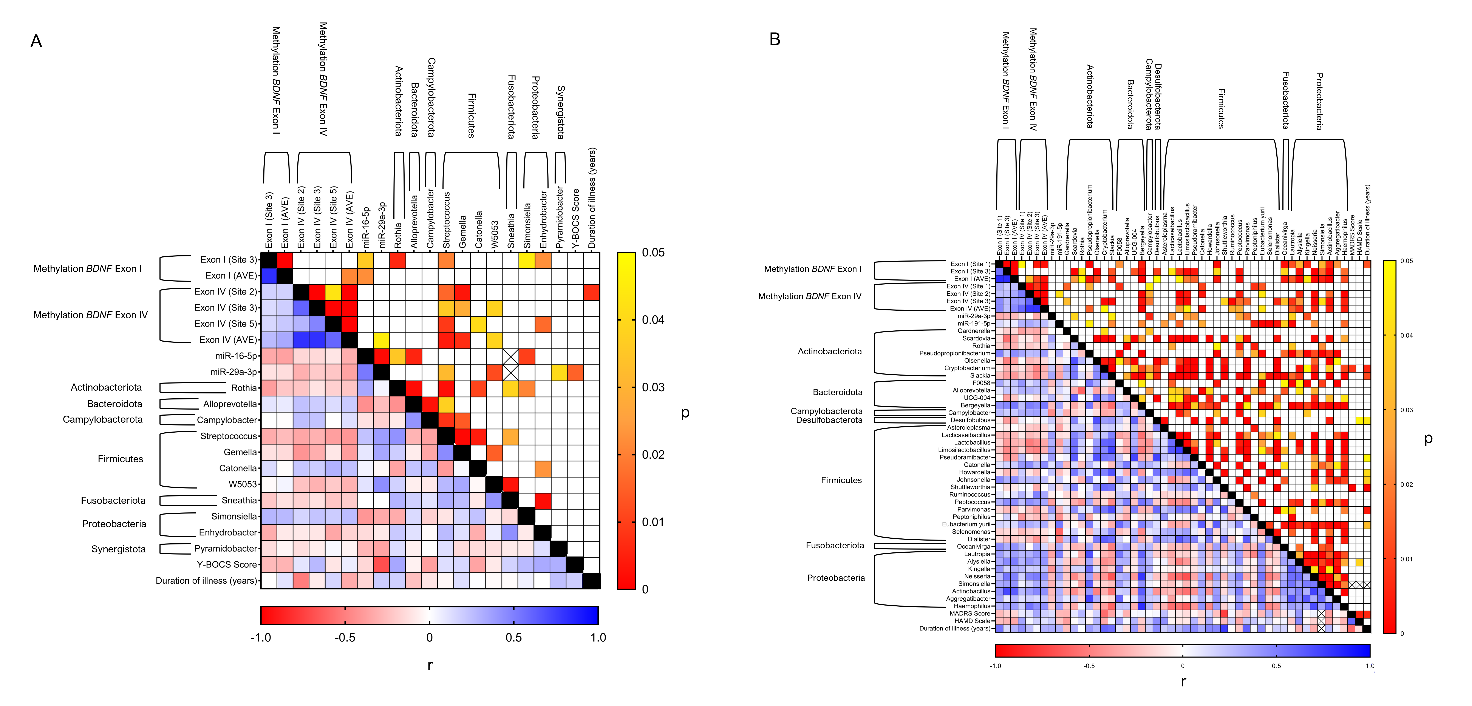

**Supplementary Figure 11.** Correlation analysis between BDNF methylation levels, miRNAs, microbiota abundance (genera) and clinical data in saliva from both CTRL and OCD individuals (**A**) and from CTRL and MDD patients (**B**). Heat maps representing the correlation analysis between BDNF methylation levels, selectively altered miRNAs, genera abundance and clinical data. Cells filled in blue to red gradient of the heat maps represent Spearman’s r; cells filled in yellow to red gradient represent p values (empty cells represent p values greater than 0.05).

*Data correlation in OCD*. Alterations in salivary microbiota significantly correlated with both epigenetic mark and miRNAs expression but not with OCD clinical parameters (Supplementary Figure 6A). In detail:

**Actinobacteriota**

- *Rothia* inversely correlated with methylation level at *BDNF* exon I, site 3 (r = - 0.382, *p* = 0.006) and directly correlated with in *miR-16-5p* expression (r = 0.343, *p* = 0.035).

**Bacteroidota**

- *Alloprevotella* resulted to inversive correlate with *miR-16-5p* (r = - 0.441, *p* = 0.006).

**Firmicutes**

- *Streptococcus* inversely correlated with methylation at *BDNF* exon I, site 3 (r = - 0.327, *p* = 0.020), at *BDNF* exon IV, site 2 (r = - 0.350, *p* = 0.016), site 3 (r = - 0.307, *p* = 0.036), site 5 (r = - 0.378, *p* = 0.009) as well as with the average of all 6 CpG sites (AVE) (r = - 0.413, *p* = 0.004) and positively correlated with *miR-29a-3p* (r = 0.380, *p* = 0.032).
- *Gemella* inversely correlated with *BDNF* exon IV, site 2 (r = - 0.421, p = 0.003), site 3 (r = - 0.320, *p* = 0.028), AVE (r = - 0.388, *p* = 0.007).
- *Catonella* positively correlated with *BDNF* exon IV, site 5 (r = 0.300, *p* = 0.041).
- *W5053* negatively correlated with *BDNF* exon IV, site 3 (r = - 0.303, *p* = 0.038), AVE (r = - 0.303, *p* = 0.039) and directly correlated with *miR-29a-3p* expression (r = 0.440, *p* = 0.012).

**Proteobacteria**

- *Simonsiella* positively correlated with *BDNF* exon I, site 3 (r = 0.284, *p* = 0.045) and negatively correlated with *miR-16-5p* (r = - 0.414, *p* = 0.010).
- *Enhydrobacter* inversely correlated with *BDNF* exon I, site 3 (r = - 0.322, *p* = 0.022), with *BDNF* exon IV, site 5 (r = - 0.348, *p* = 0.017).

**Synergistota**

- *Pyramidobacter* negatively correlated with *miR-29a-3p* (r = - 0.365, *p* = 0.040).

Moreover:

- *BDNF* exon I, site 3 and AVE inversely correlated with *miR-16-5p* (r = - 0.341, *p* = 0.036 and r = - 0.370, *p* = 0.022, respectively).
- *BDNF* exon IV, AVE resulted to be inversive correlated with *miR-29a-3p* (r = - 0.362, *p* = 0.045).
- *BDNF* exon IV, site 2 inversely correlated with the duration of illness (r = - 0.512, *p* = 0.007).
- *miR-29a-3p* inversely correlated with Y-BOCS score (r = - 0.692, *p* = 0.015).

*Data correlation in MDD*. Salivary microbiota significantly correlated with DNA methylation level at *BDNF* gene, with miRNAs and with clinical parameters belonging to MDD patients (Supplementary Figure 6B), specifically:

**Actinobacteriota**

- *Gardnerella* inversely correlated with DNA methylation at *BDNF* exon IV, site 1 (r = - 0.376, *p* = 0.015) and site 3 (r = - 0.335, *p* = 0.032).
- *Scardovia* inversely correlated with *BDNF* exon I, site 3 (r = - 0.567, *p* < 0.0001) and AVE (r = - 0.338, *p* = 0.025) and directly correlated with *miR-29a-3p* (r = 0.350, *p* = 0.039).
- *Rothia* inversely correlated *BDNF* exon I, site 3 (r = - 0.310, *p* = 0.043) and AVE (r = - 0.298, *p* = 0.049).
- *Pseudopropionibacterium* directly correlated with *BDNF* exon I, site 1 (r = 0.497, *p* = 0.001), AVE (r = 0.441, *p* = 0.003), with *BDNF* exon IV, site 1 (r = 0.342, *p* = 0.029), AVE (r = 0.377, *p* = 0.015) and with *miR-191-5p* (r = 0.318, *p* = 0.046).
- *Olsenella* resulted to inversive correlate with *BDNF* exon I, site 3 (r = - 0.500, *p* = 0.001) and AVE (r = - 0.379, *p* = 0.011).
- *Cryptobacterium* inversely correlated with BDNF exon IV, site 3 (r = - 0.482, *p* = 0.001) and AVE (r = - 0.310, *p* = 0.049). Also, resulted to be inversely correlated with MADRS score (r = - 0.576, *p* = 0.008) and positively correlated with the duration of illness (r = 0.593, *p* = 0.006).
- *Slackia* showed an inversive correlation with *BDNF* exon I, site 3 (r = - 0.374, *p* = 0.014) and AVE (r = - 0.400, *p* = 0.007), and *BDNF* exon IV, site 3 (r = - 0.480, *p* = 0.001). Additionally, was directly correlated with *miR-29a-3p* (r = 0.340, *p* = 0.045) and inversely with *miR-191-5p* (r = - 0.362, *p* = 0.022). Resulted also to positively correlate with duration of illness (r = 0.555, *p* = 0.011).

**Bacteroidota**

- *F0058* directly correlated with *BDNF* exon I, site 1 (r = 0.342, *p* = 0.023), *BDNF* exon IV, site 1 (r = 0.411, *p* = 0.008).
- *UCG-004* inversely correlated with *BDNF* exon I, site 3 (r = - 0.376, *p* = 0.013).
- *Bergeyella* positively correlated with DNA methylation at *BDNF* gene in both promoter regions. In particular, exon I, site 1 (r = 0.417, *p* = 0.005), site 3 (r = 0.463, *p* = 0.002), AVE (r = 0.525, *p* < 0.0001), exon IV, site 2 (r = 0.500, *p* = 0.001), site 3 (r = 0.455, *p* = 0.003), AVE (r = 0.521, *p* < 0.0001). Moreover, positively correlated with *miR-191-5p* (r = 0.361, *p* = 0.022).

**Campylobacterota**

- *Campylobacter* positively correlated with *BDNF* exon IV, site 1 (r = 0.313, *p* = 0.046), site 2 (r = 0.342, *p* = 0.028), site 3 (r = 0.366, *p* = 0.018) and inversely correlated with *miR-29a-3p* (r = - 0.458, *p* = 0.006).

**Desulfobacterota**

- *Desulfobulbus* inversely correlated with *BDNF* exon I, site 3 (r = - 0.391, *p* = 0.010) and AVE (r = - 0.298, *p* = 0.049). Also, positively correlated with HAMD scale (r = 0.449, *p* = 0.047) and duration of illness (r = 0.454, *p* = 0.044).

**Firmicutes**

- *Asteroleplasma* inversely correlated with *miR-191-5p* (r = - 0.355, *p* = 0.025).
- *Lacticaseibacillus* inversely correlated with *BDNF* exon I, site 3 (r = - 0.310, *p* = 0.043) and positively correlated with *miR-29a-3p* (r = 0.380, *p* = 0.024).
- *Lactobacillus* inversely correlated with *BDNF* exon I, site 3 (r = - 0.396, *p* = 0.009), AVE (r = - 0.301, *p* = 0.047), with *BDNF* exon IV, site 2 (r = - 0.520, *p* < 0.0001), site 3 (r = - 0.450, *p* = 0.003), AVE (r = - 0.345, *p* = 0.027) and, also, inversely correlated with *miR-191-5p* (r = - 0.379, *p* = 0.016).
- *Limosilactobacillus* inversely correlated with *BDNF* exon I, site 3 (r = - 0.472, *p* = 0.001), AVE (r = - 0.351, *p* = 0.020), with *BDNF* exon IV, site 2 (r = - 0.443, *p* = 0.004), site 3 (r = - 0.497, *p* = 0.001), AVE (r = - 0.313, *p* = 0.046) and resulted to correlate positively with *miR-29a-3p* (r = 0.374, *p* = 0.027).
- *Pseudoramibacter* inversely correlated with *BDNF* exon I, site 3 (r = - 0.436, *p* = 0.003), AVE (r = - 0.363, *p* = 0.015) and positively with *miR-29a-3p* (r = 0.382, *p* = 0.023), moreover, positively correlated with duration of illness (r = 0.448, *p* = 0.048).
- *Catonella* correlated positively with *BDNF* exon IV, site 1 (r = 0.398, *p* = 0.010), site 2 (r = 0.450, *p* = 0.003) and with *BDNF* exon IV AVE (r = 0.444, *p* = 0.004). Resulted to positively correlate with *miR-191-5p* (r = 0.348, *p* = 0.028).
- *Howardella* positively correlated with duration of illness (r = 0.461, *p* = 0.041).
- *Johnsonella* positively correlated with *BDNF* exon I, site 1 (r = 0.462, *p* = 0.002), site 3 (r = 0.314, *p* = 0.040), and AVE (r = 0.431, *p* = 0.003) and also correlated with the duration of illness (r = 0.468, *p* = 0.037).
- *Shuttleworthia* inversely correlated with *BDNF* exon IV, site 3 (r = - 0.325, *p* = 0.038).
- *Ruminococcus* positively correlated with *BDNF* exon IV, site 3 (r = 0.492, *p* = 0.001), and AVE (r = 0.340, *p* = 0.030).
- *Peptococcus* positively correlated with *BDNF* exon I, site 3(r = 0.438, p = 0.003), AVE (r = 0.442, *p* = 0.003), with *BDNF* exon IV, site 2 (r = 0.432, *p* = 0.005), site 3 (r = 0.370, *p* = 0.017) and AVE (r = 0.342, *p* = 0.029) and inversely correlated with *miR-29a-3p* (r = - 0.373, *p* = 0.027).
- *Parvimonas* inversely correlated with *BDNF* exon IV, site 3 (r = - 0.376, *p* = 0.015), directly with *miR-29a-3p* (r = 0.399, *p* = 0.018) and also directly correlated with the duration of illness (r = 0.619, *p* = 0.004).
- *Peptoniphilus* inversely correlated with *BDNF* exon IV, site 1 (r = - 0.411, *p* = 0.008) and with *miR-191-5p* (r = - 0.435, *p* = 0.005).
- *Eubacterium yurii* positively correlated with *BDNF* exon IV, site 1 (r = 0.383, *p* = 0.013), site 2 (r = 0.317, *p* = 0.044), AVE (r = 0.319, *p* = 0.042), *miR-191-5p* (r = 0.528, *p* < 0.0001) and with the duration of illness (r = 0.479, *p* = 0.033).
- *Selenomonas* inversely correlated with *miR-191-5p* (r = - 0.604, *p* < 0.0001).
- *Dialister* inversely correlated with *BDNF* exon IV, site 3 (r = - 0.421, *p* = 0.006), *miR-191-5p* (r = - 0.465, *p* = 0.002).

**Fusobacteriota**

- *Oceanivirga* directly correlated with *BDNF* exon I, site 1 (r = 0.447, *p* = 0.002), site 3 (r = 0.316, *p* = 0.039), AVE (r = 0.427, *p* = 0.004), with *BDNF* exon IV, site 3 (r = 0.333, *p* = 0.034), *miR-191-5p* (r = 0.332, *p* = 0.037)

**Proteobacteria**

- *Lautropia* directly correlated with *BDNF* exon I, site 1 (r = 0.374, *p* = 0.012), AVE (r = 0.369, *p* = 0.014), with *BDNF* exon IV, site 3 (r = 0.408, *p* = 0.008), AVE (r = 0.377, *p* = 0.015), with *miR-191-5p* (r = 0.494, *p* = 0.001) and inversely with *miR-29a-3p* (r = - 0.353, *p* = 0.037).
- *Alysiella* directly correlated with *BDNF* exon I, site 1 (r = 0.327, *p* = 0.030), site 3 (r = 0.454, *p* = 0.002), AVE (r = 0.507, *p* < 0.001), with *BDNF* exon IV, site 2 (r = 0.420, *p* = 0.006), site 3 (r = 0.353, *p* = 0.024), AVE (r = 0.414, *p* = 0.007) and also with HAMD scale (r = 0.466, *p* = 0.038).
- *Kingella* directly correlated with *BDNF* exon I, site 1 (r = 0.411, *p* = 0.006) and AVE (r = 0.306, *p* = 0.044) and inversely with *miR-29a-3p* (r = - 0.419, *p* = 0.012).
- *Neisseria* directly correlated with *BDNF* exon I, site 3 (r = 0.541, *p* < 0.001), AVE (r = 0.489, *p* = 0.001), with *BDNF* exon IV, site 2 (r = 0.476, *p* = 0.002), site 3 (r = 0.513, *p* = 0.001), AVE (r = 0.420, *p* = 0.006) and with *miR-191-5p* (r = 0.359, *p* = 0.023).
- *Simonsiella* directly correlated with *BDNF* exon I, site 1 (r = 0.326, *p* = 0.031), site 3 (r = 0.364, *p* = 0.017), AVE (r = 0.418, *p* = 0.005) and with *BDNF* exon IV, site 3 (r = 0.321, *p* = 0.041).
- *Actinobacillus* positively correlated with *BDNF* exon I, site 1 (r = 0.413, *p* = 0.005), site 3 (r = 0.710, *p* < 0.0001), AVE (r = 0.635, *p* < 0.0001) and with *BDNF* exon IV, site 2 (r = 0.371, *p* = 0.017), site 3 (r = 0.459, *p* = 0.003), AVE (r = 0.433, *p* = 0.005).
- *Aggregatibacter* directly correlated with *BDNF* exon I, site 1 (r = 0.341, *p* = 0.024).
- *Haemophilus* directly correlated with *BDNF* exon I, site 3 (r = 0.362, *p* = 0.017), AVE (r = 0.378, *p* = 0.011), with *BDNF* exon IV, site 2 (r = 0.543, *p* < 0.001), site 3 (r = 0.429, p = 0.005), AVE (r = 0.436, *p* = 0.004) and with *miR-191-5p* (r = 0.377, *p* = 0.017).

Moreover:

- *BDNF* exon I, site 1 positively correlated with the duration of illness (r = 0.548, *p* = 0.012).
- *BDNF* exon IV, site 2 (r = 0.335, *p* = 0.043).
