## Supplementary figures and images for "DISTINCT SALIVARY MICROBIOTA PROFILES, *BDNF* DNA METHYLATION AND *MIR-16-5P*, *MIR-29A-3P*, *MIR-191-5P* ALTERATIONS IN OBSESSIVE-COMPULSIVE DISORDER AND MAJOR DEPRESSIVE DISORDER"

### Figure S1.tiff

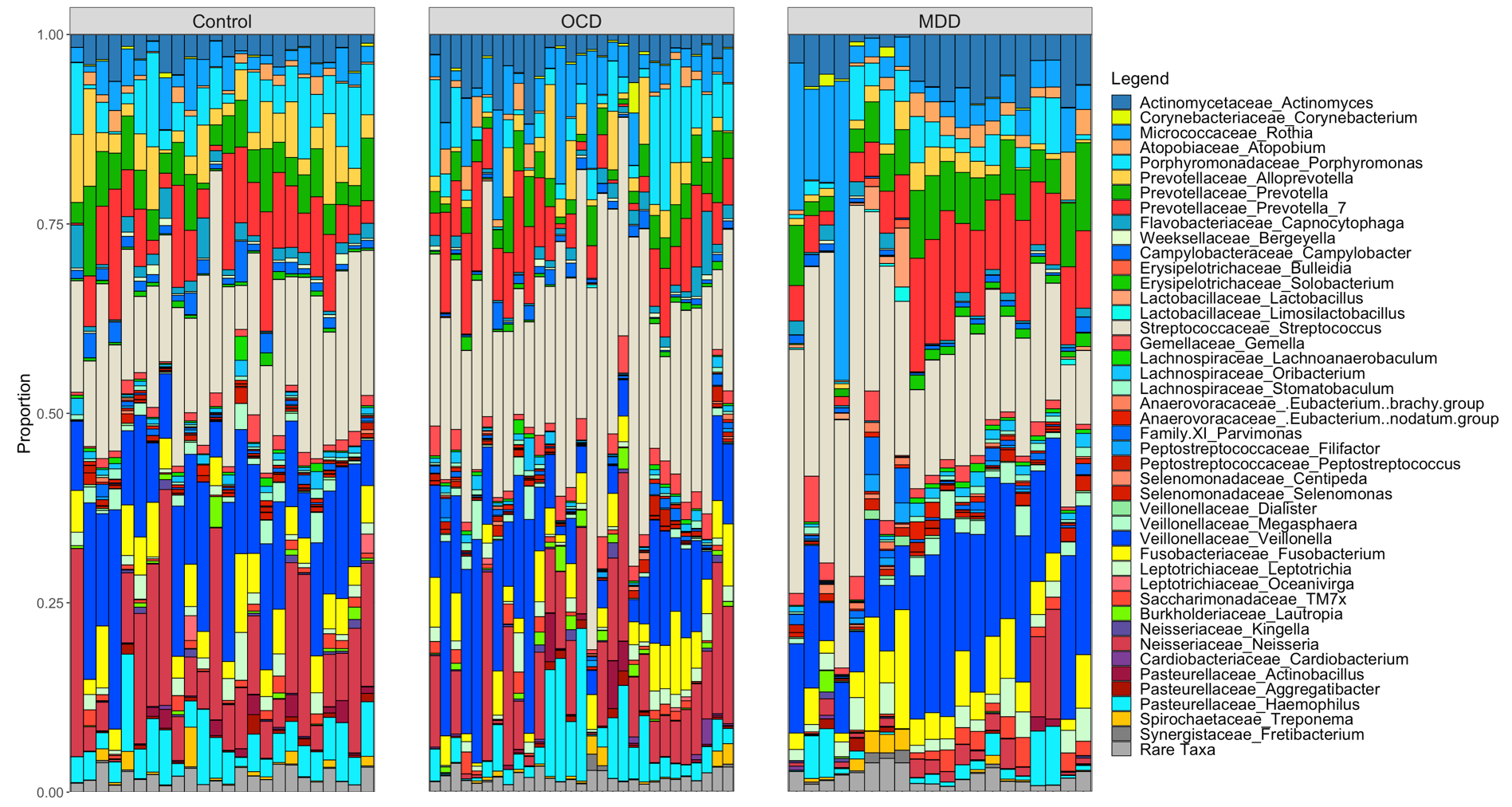

### Figure S2.tiff

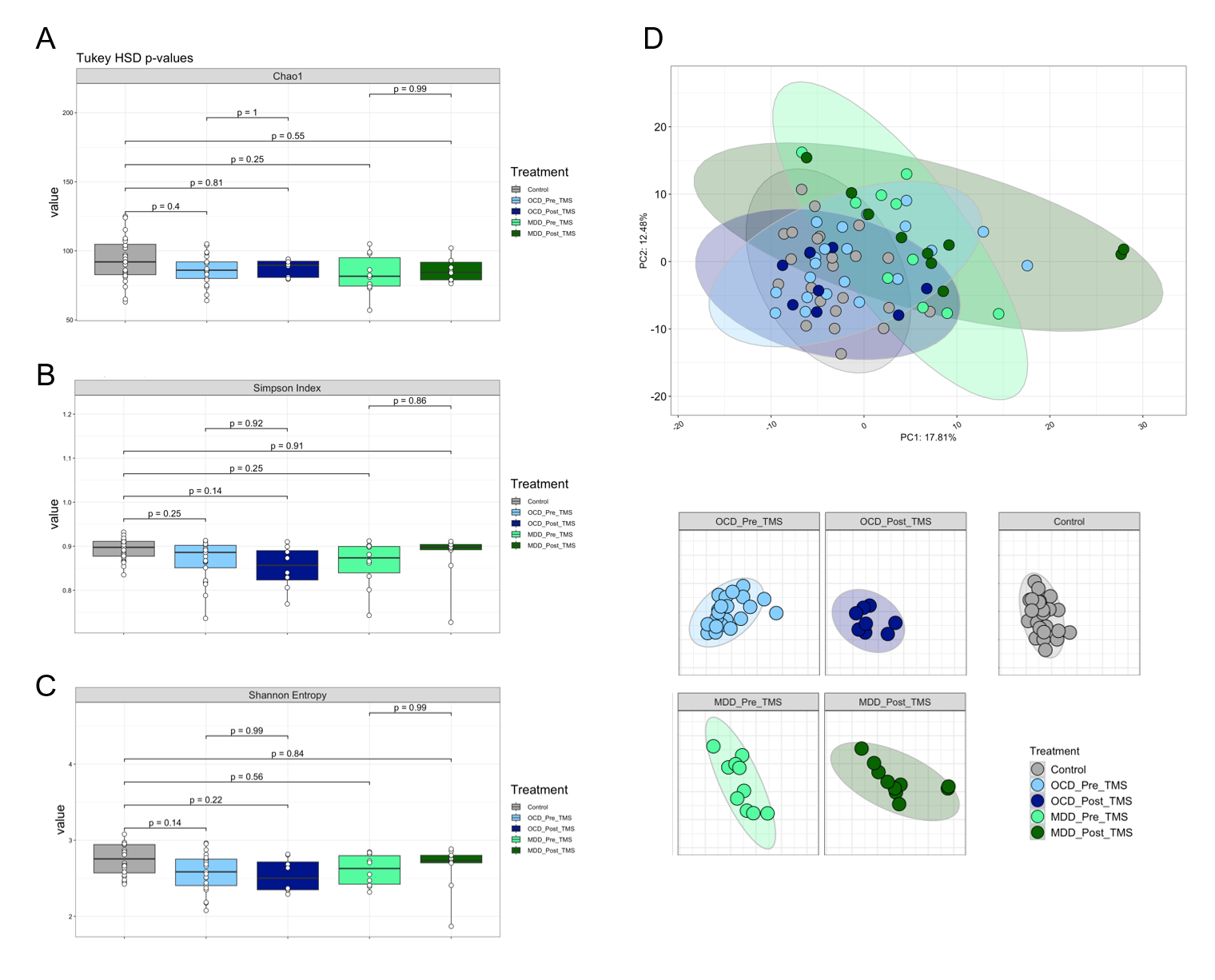

### Figure S3.tiff

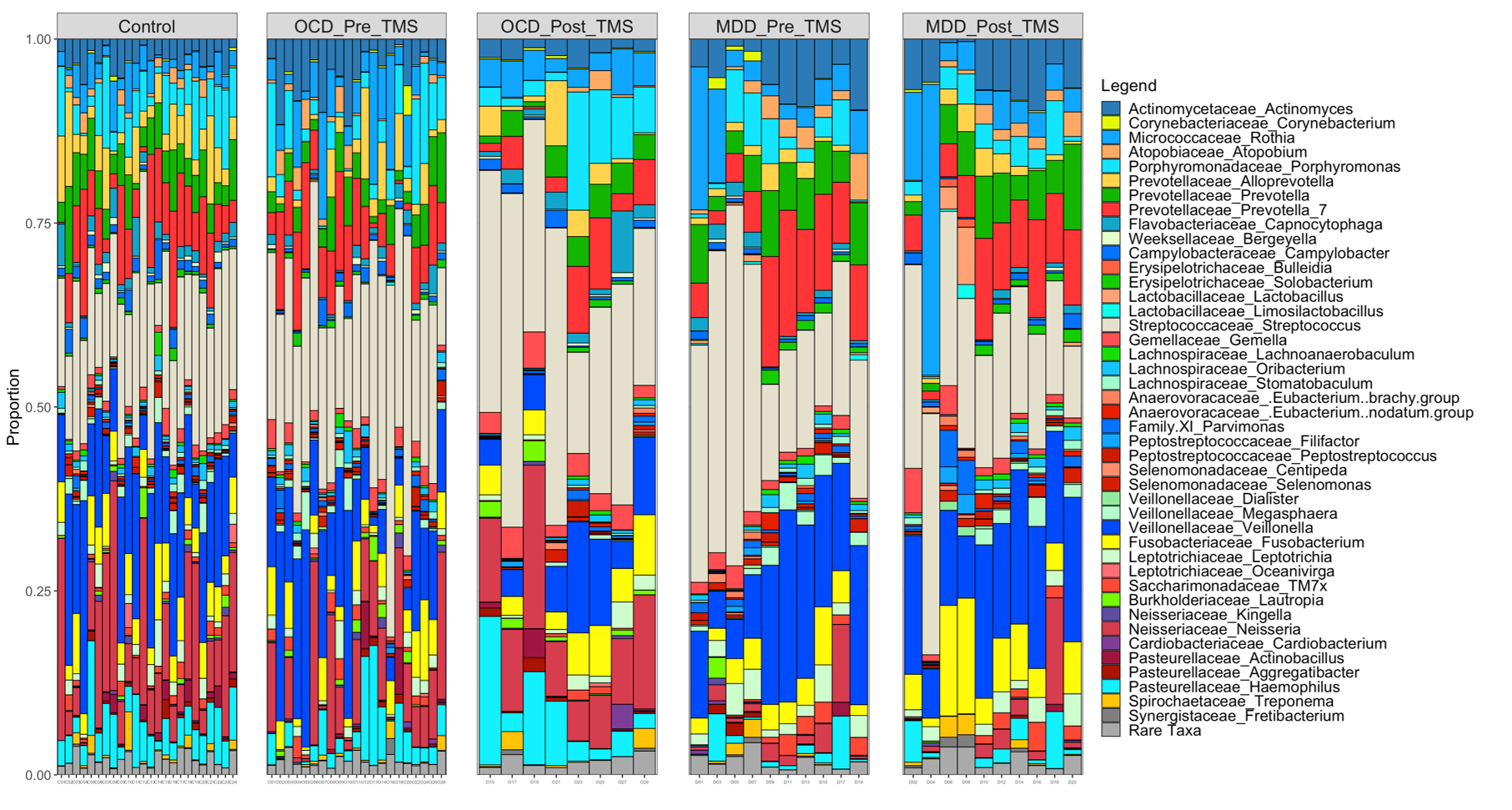

### Figure S4.tiff

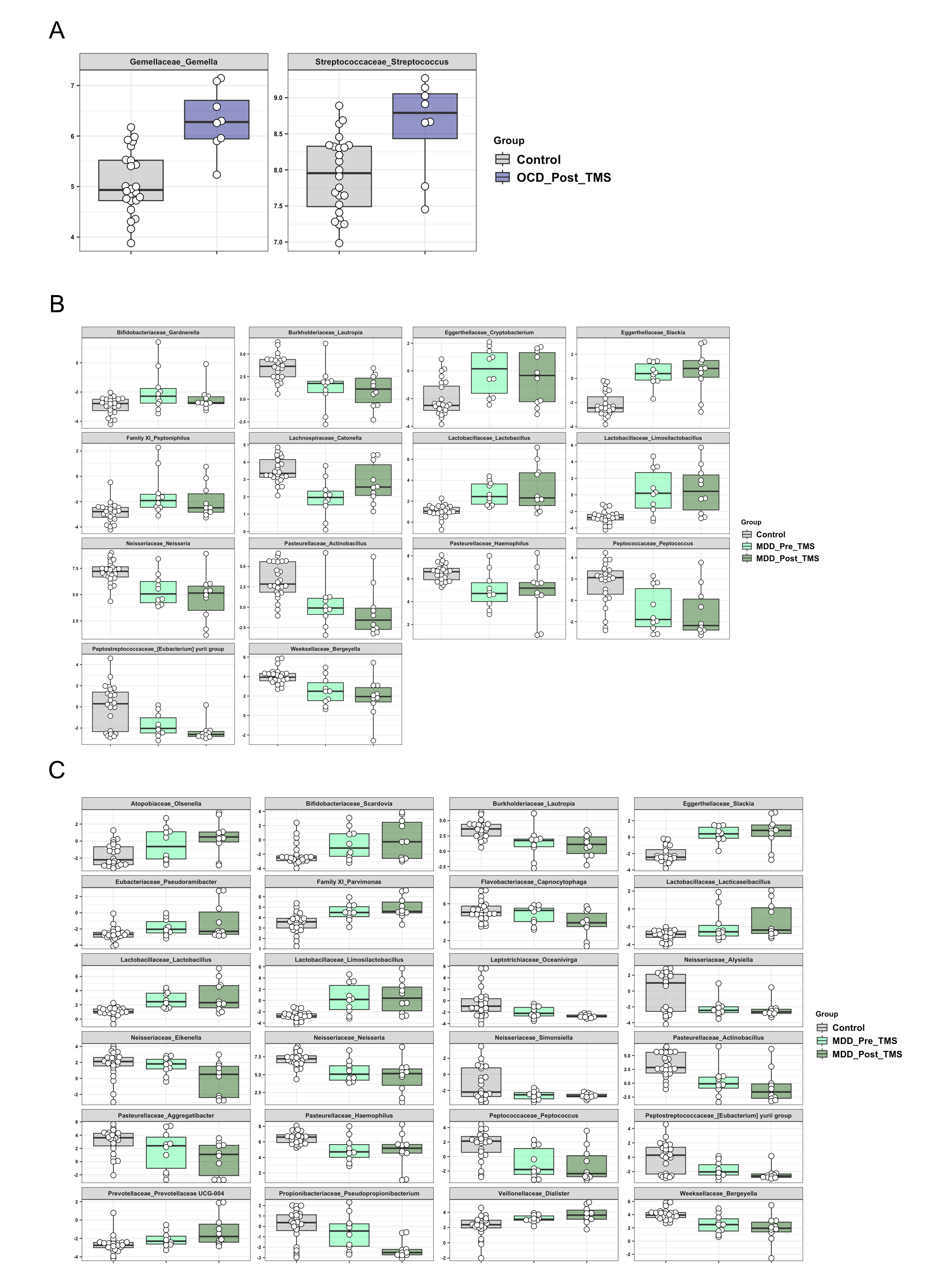

### Figure S5.tiff

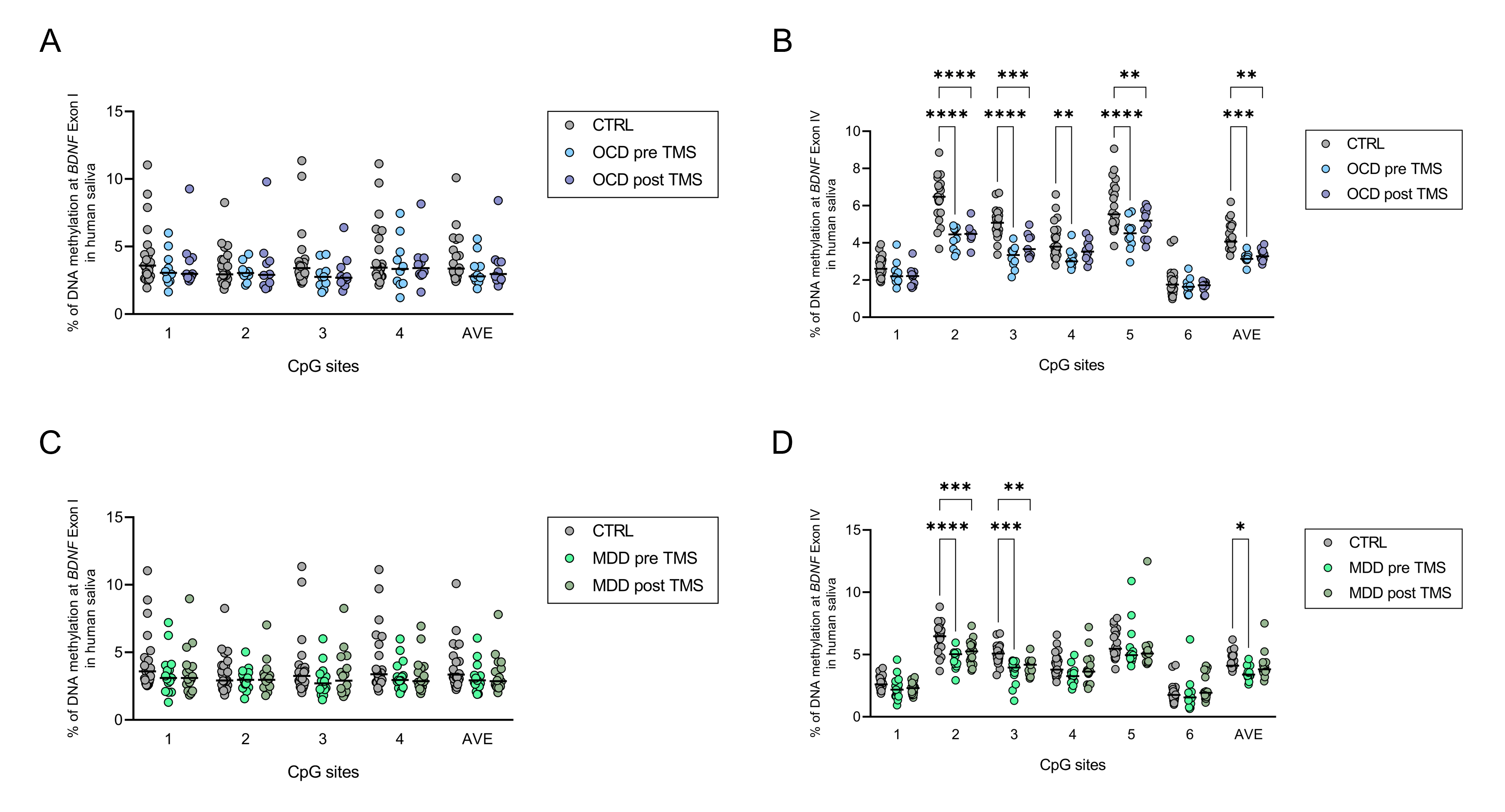

### Figure S6.tiff

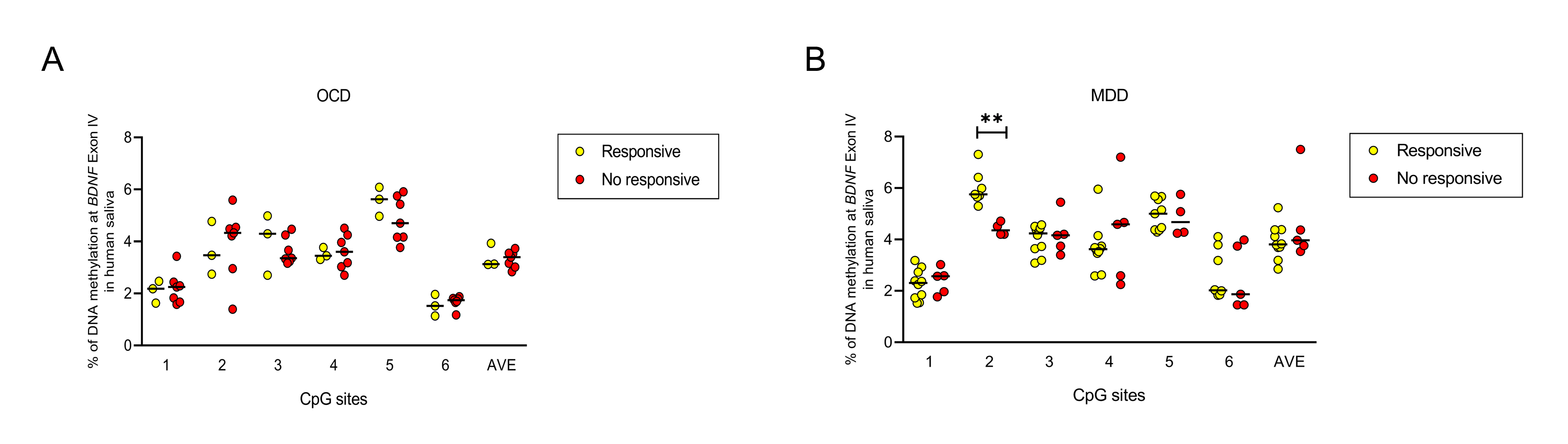

### Figure S7.tiff

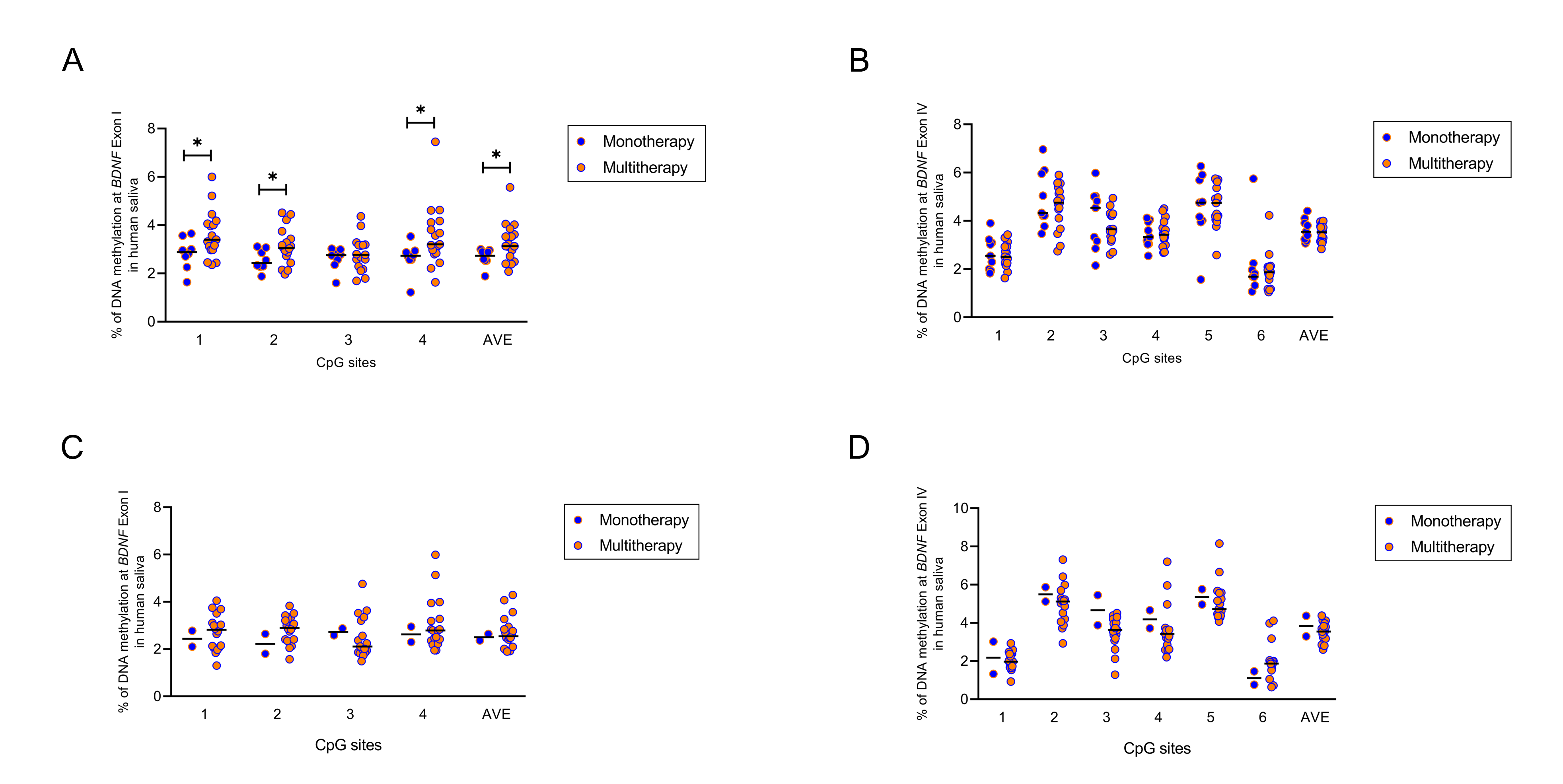

### Figure S8.tiff

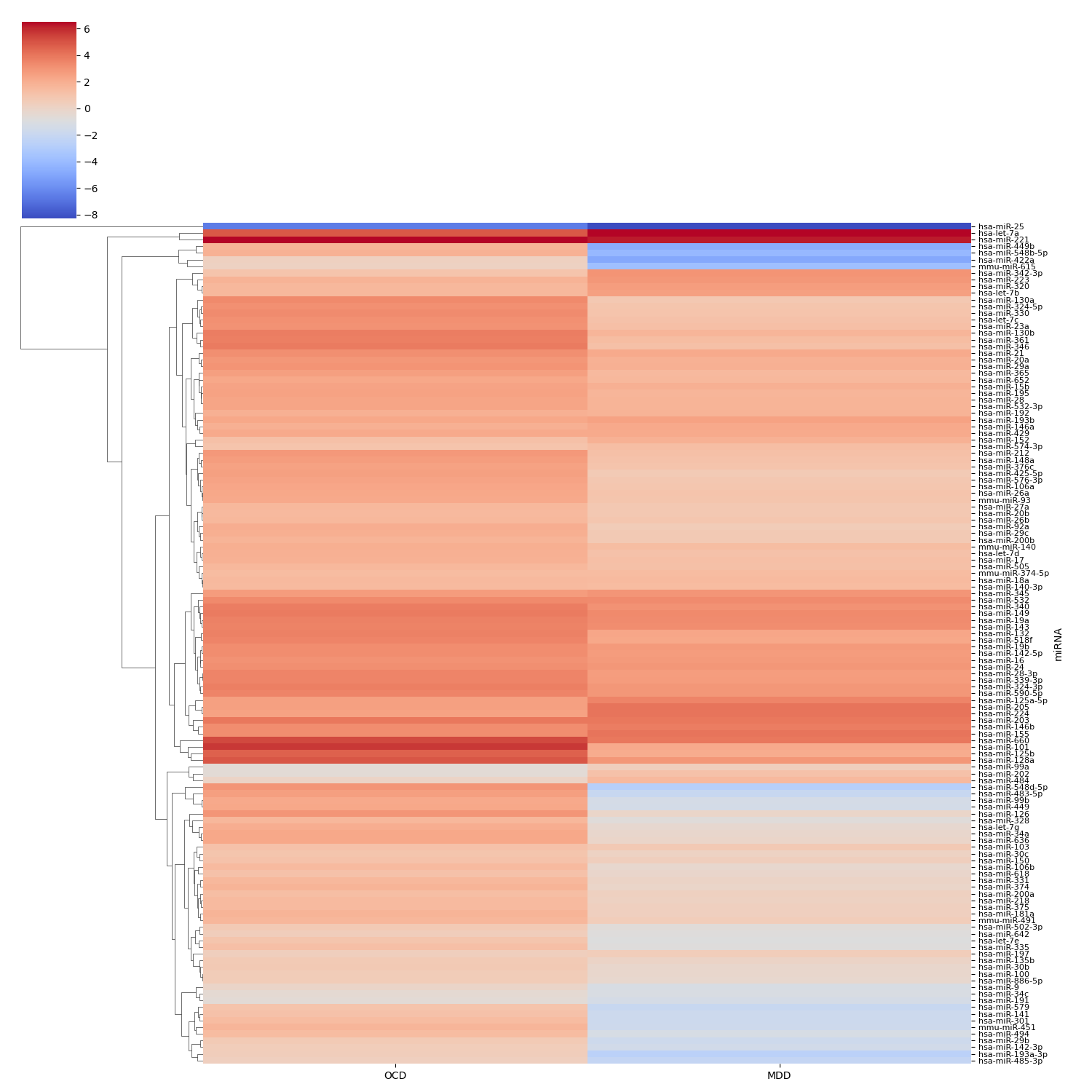

### Figure S9.tiff

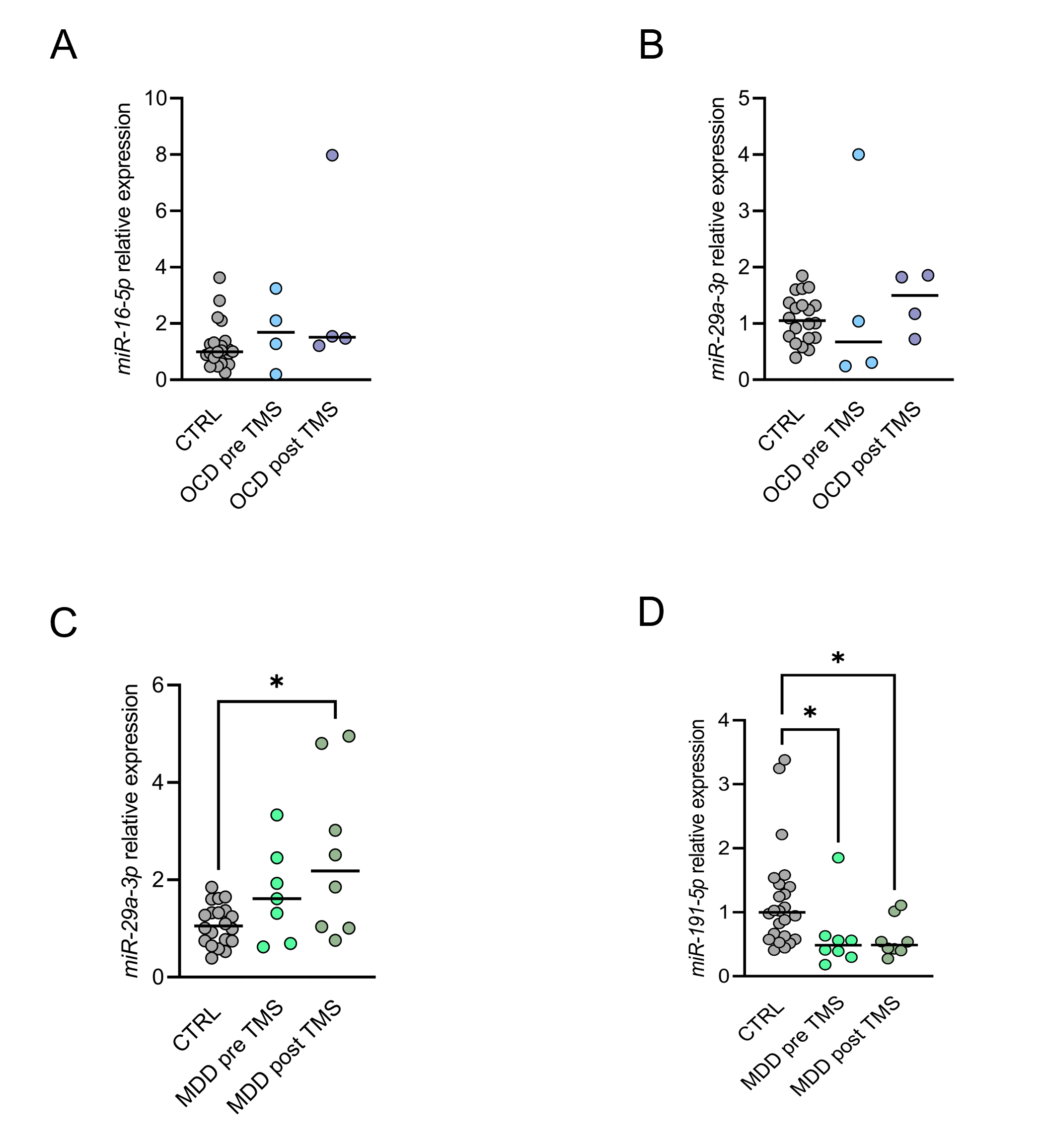

### Figure S10.tiff

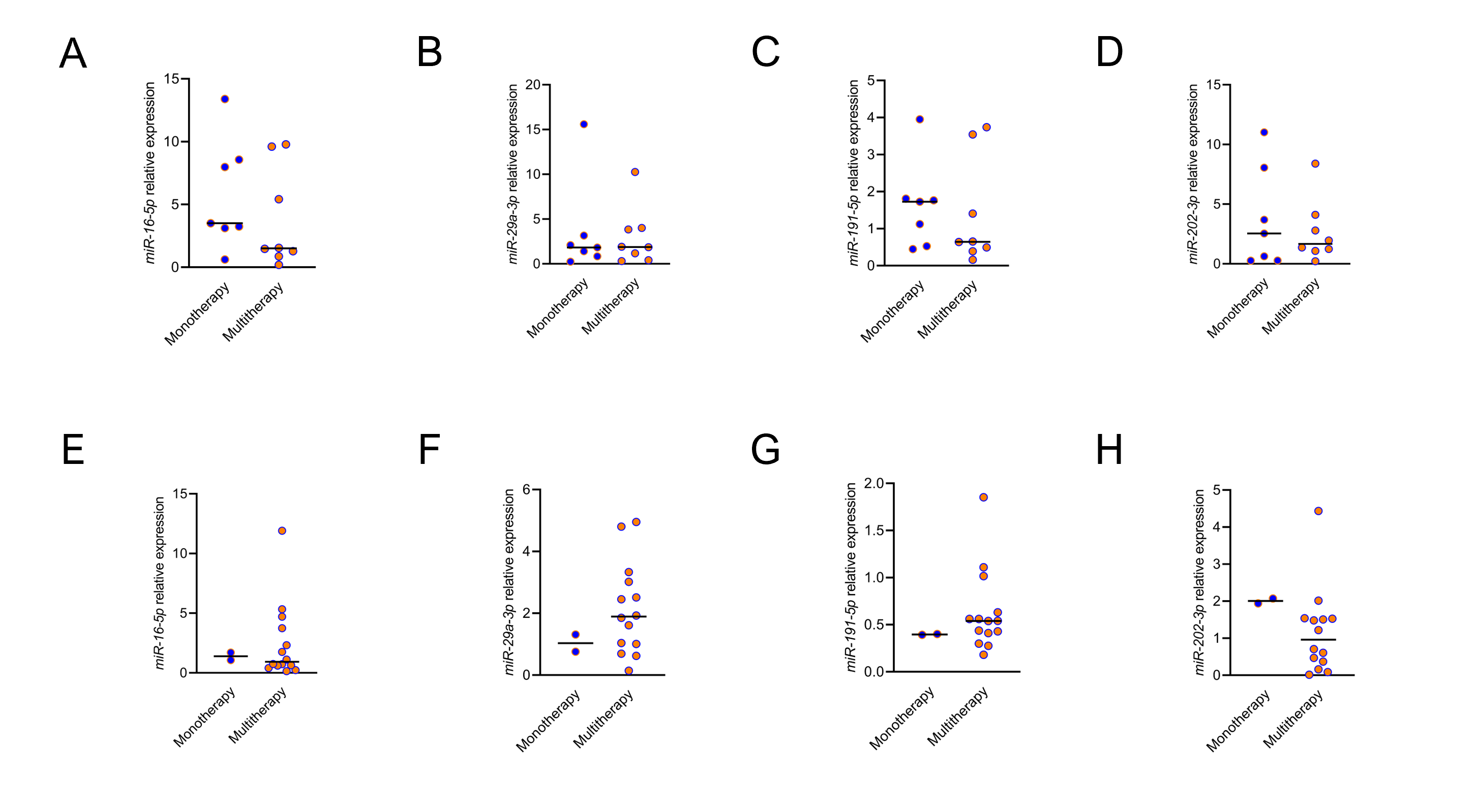
